# Doubling Time of the COVID-19 Epidemic by Chinese Province

**DOI:** 10.1101/2020.02.05.20020750

**Authors:** Kamalich Muniz-Rodriguez, Gerardo Chowell, Chi-Hin Cheung, Dongyu Jia, Po-Ying Lai, Yiseul Lee, Manyun Liu, Sylvia K. Ofori, Kimberlyn M. Roosa, Lone Simonsen, Cecile Viboud, Isaac Chun-Hai Fung

**Affiliations:** Georgia Southern University, Statesboro, GA, USA; Georgia State University, Atlanta, GA, USA; Independent researcher; Boston University, Boston, MA, USA; Roskilde University, Roskilde, Denmark; The National Institutes of Health, Bethesda, MD, USA

**Keywords:** China, Coronavirus, COVID-19, Disease Transmission, Infectious, Epidemiology, Respiratory Tract Infections

## Abstract

COVID-19 epidemic doubling time by Chinese province was increasing from January 20 through February 9, 2020. The harmonic mean of the arithmetic mean doubling time estimates ranged from 1.4 (Hunan, 95% CI, 1.2-2.0) to 3.1 (Xinjiang, 95% CI, 2.1-4.8), with an estimate of 2.5 days (95% CI, 2.4-2.6) for Hubei.

## Main text

Our ability to estimate the basic reproduction number of emerging infectious diseases is often hindered by the paucity of information about the epidemiological characteristics and transmission mechanisms of new pathogens (1). Alternative metrics could synthesize real-time information about the extent to which the epidemic is expanding over time. Such metrics would be particularly useful if they rely on minimal and routinely collected data that capture the trajectory of an outbreak (2).

Epidemic doubling times characterize the sequence of intervals at which the cumulative incidence doubles (3). If an epidemic is growing exponentially with a constant growth rate *r*, the doubling time remains constant and equals to (ln 2)/*r*. An increase in the doubling time indicates a slowdown in transmission if the underlying reporting rate remains unchanged (Technical Appendix) (4).

Here we analyzed by province the number of times COVID-19 cumulative incidence doubled and the evolution of the doubling times in mainland China (5), from January 20 (when nationwide reporting began) through February 9, 2020. Province-level daily cumulative incidence data were retrieved from provincial health commissions’ websites. Two sensitivity analyses based on a longer and a shorter time period respectively were conducted (Technical Appendix). Tibet was excluded from further analysis because there had only been one case reported during the study period.

From January 20 through February 9, the harmonic mean of the arithmetic means of the doubling times estimated from cumulative incidence ranged from 1.4 (95% CI, 1.2, 2.0) days (Hunan) to 3.1 (95%CI, 2.1, 4.8) days (Xinjiang). In Hubei, it was estimated as 2.5 (95% CI, 2.4, 2.6) days. The cumulative incidence doubled 6 times in Hubei. The harmonic mean of the arithmetic means of doubling times in all of mainland China except Hubei was 1.8 (95% CI, 1.5, 2.3) days. Provinces with a harmonic mean of the arithmetic means of doubling times <2d included Fujian, Guangxi, Hebei, Heilongjiang, Henan, Hunan, Jiangxi, Shandong, Sichuan, and Zhejiang (Figures 1 and S1).

**Figure 1.**
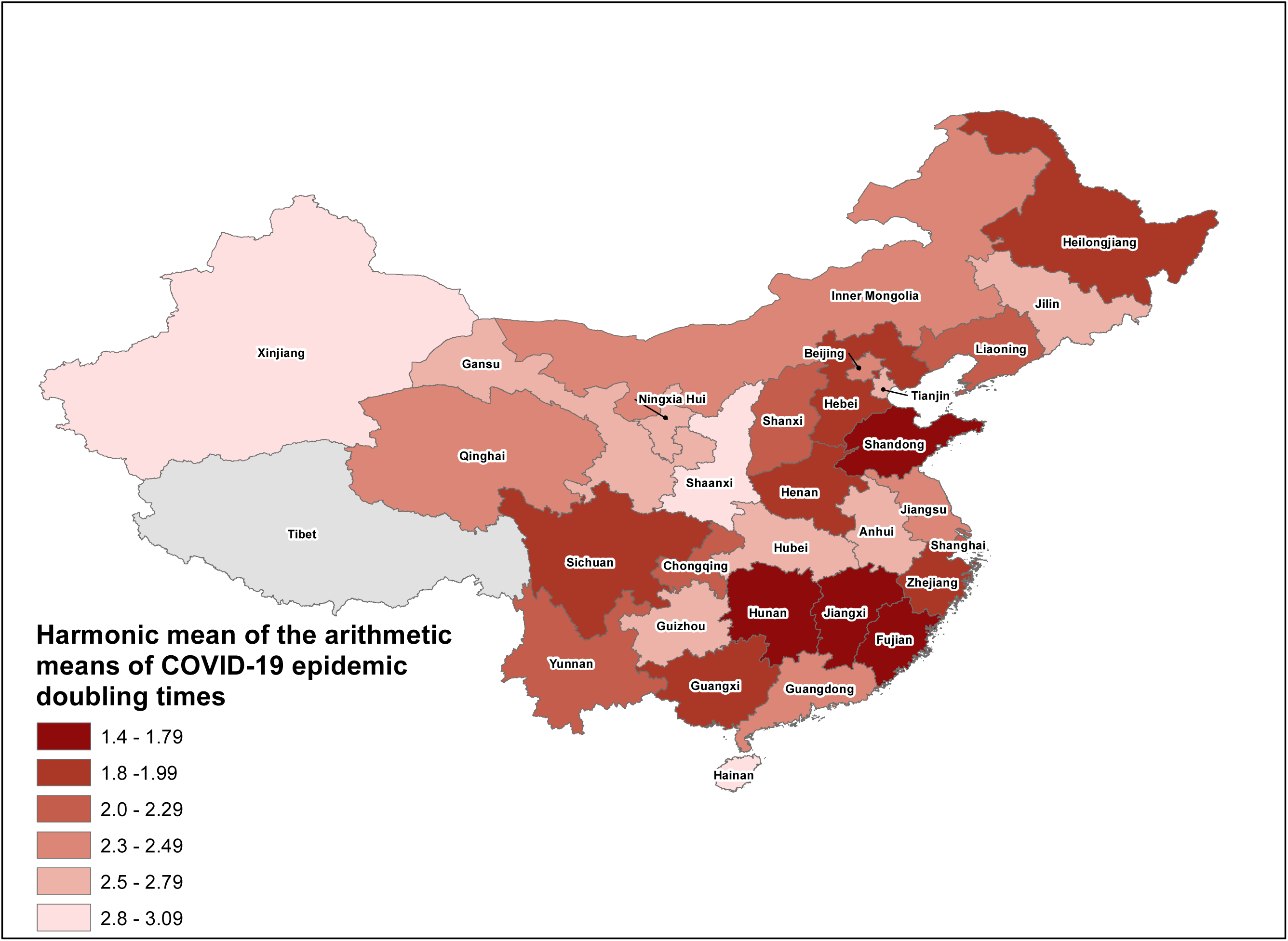
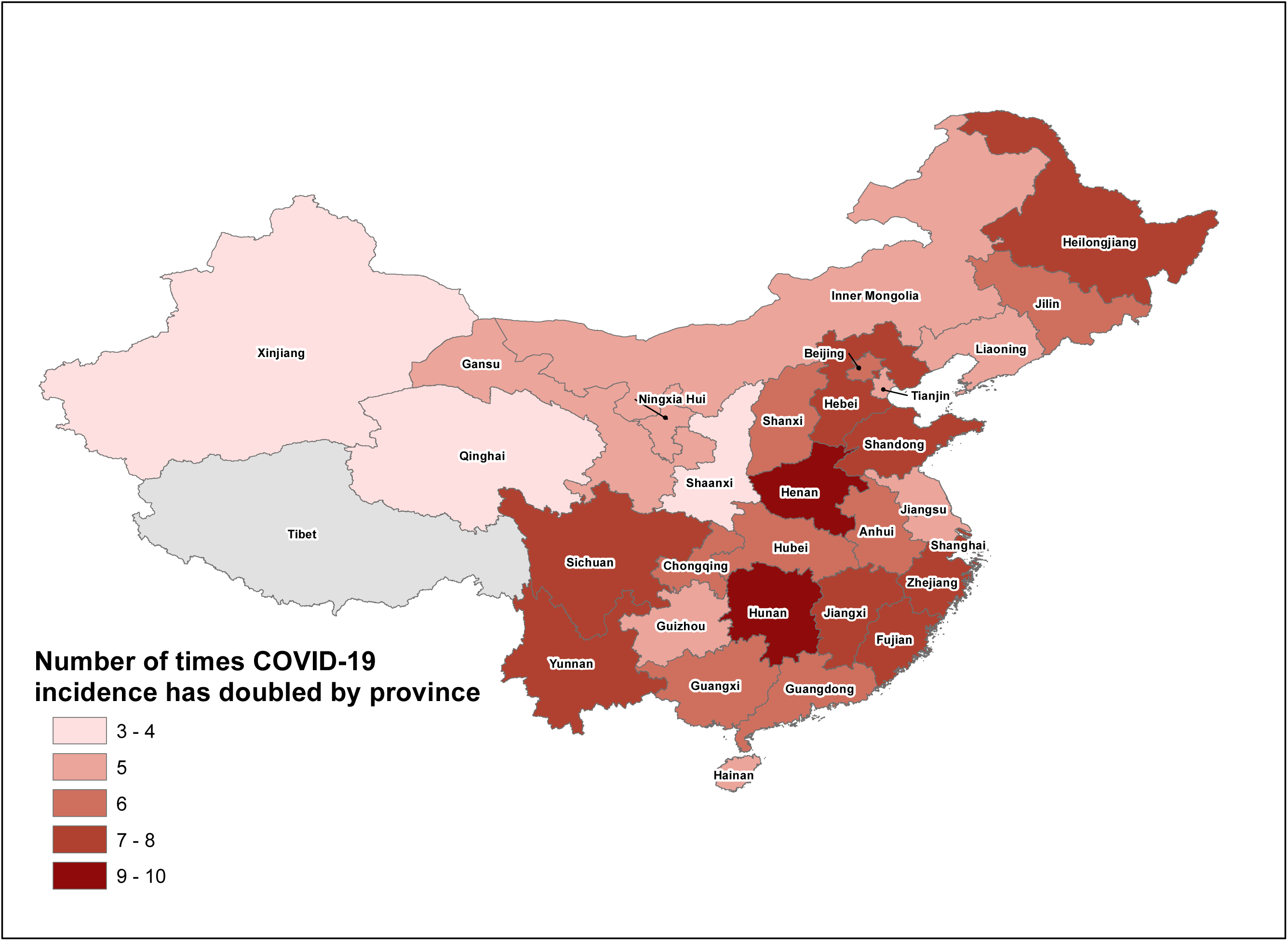
The map of the harmonic mean of the arithmetic means of doubling time estimates (Panel A) and the number of times the COVID-19 epidemic cumulative incidence has doubled (Panel B) by province in mainland China, from January 20 through February 9, 2020.

As the epidemic progressed, it took longer for the cumulative incidence in mainland China (except Hubei) to double itself, which indicated an overall sub-exponential growth pattern outside Hubei (Figures S1, S2). In Hubei, the doubling time decreased and then increased. A gradual increase in the doubling time coincided with the social distancing measures and intra-and-inter-provincial travel restrictions imposed across China since the implementation of quarantine of Wuhan on January 23 (6).

Our estimates of doubling times are shorter than prior estimates of 7.4 days (95% CI, 4.2-14) (5), 6.4 days (95% CrI, 5.8-7.1) (7), and 7.1 days (95% CI, 3.0-20.5) (8) respectively. Li et al. covered cases reported by January 22 (5). Wu et al. statistically inferred case counts in Wuhan by internationally exported cases as of January 25 (7). Volz et al. identified a common viral ancestor on December 8, 2019 using Bayesian phylogenetic analysis and fitted an exponential growth model to provide the epidemic growth rate (8). Our estimates are based on cumulative confirmed case count by reporting date by province from January 20 through February 9.

Our study is subject to limitations, including underreporting of cases (9). One reason for underreporting is underdiagnosis, due to lack of diagnostic tests, healthcare workers, and other resources. Further, underreporting is likely heterogeneous across provinces. As long as reporting remains invariant over time within the same province, the calculation of doubling times remains reliable; however, this is a strong assumption. Growing awareness of the epidemic and increasing availability of diagnostic tests might have strengthened reporting over time, which could have artificially shortened the doubling time. Nevertheless, apart from Hubei and Guangdong (first case reported on January 19), nationwide reporting only began on January 20, and at this point, Chinese authorities had openly acknowledged the magnitude and severity of the epidemic. Due to a lack of detailed case data describing incidence trends for imported and local cases, we focused our analysis on the overall trajectory of the epidemic without adjusting for the role of imported cases on the local transmission dynamics. Indeed, it is likely that the proportion of imported cases was significant for provinces that only reported a few cases; their short doubling times in the study period could simply reflect rapid detection of imported cases. However, with the data until February 9, only two provinces had a cumulative case count of <40 (Table S1). It would be interesting to investigate the evolution of the doubling time after accounting for case importations if more detailed data becomes available.

To conclude, we observed an increasing trend in the epidemic doubling time of COVID-19 by Chinese province from January 20 through February 9, 2020. The harmonic mean of the arithmetic means of doubling times of cumulative incidence in Hubei during the study period was estimated at 2.5 (95% CI, 2.4, 2.6) days.

## Data Availability

All data analyzed is publicly available aggregated data. The data is provided in the Technical Appendix.

## Acknowledgements

GC acknowledges support from NSF grant 1414374 as part of the joint NSF-NIH-USDA Ecology and Evolution of Infectious Diseases program. ICHF acknowledges salary support from the National Center for Emerging and Zoonotic Infectious Diseases, Centers for Disease Control and Prevention (19IPA1908208). This article is not part of ICHF’s CDC-sponsored projects.

## Disclaimers

This article does not represent the official positions of the Centers for Disease Control and Prevention, the National Institutes of Health, or the United States Government.

## Author Bio

Kamalich Muniz-Rodriguez, MPH, is a doctoral student at the Jiann-Ping Hsu College of Public Health, Georgia Southern University. Her research interests include infectious disease epidemiology, digital epidemiology and disaster epidemiology.

Gerardo Chowell, PhD, is Professor of Epidemiology and Biostatistics, and Chair of the Department of Population Health Sciences at Georgia State University School of Public Health. As a mathematical epidemiologist, Prof Chowell studies the transmission dynamics of emerging infectious diseases, such as Ebola, MERS and SARS.

## Technical appendix

### Additional information on our motivation, scope and methods

#### Motivation

R_0_ is a widely used indicator of transmission potential in a totally susceptible population and is driven by the average contact rate and the mean infectious period of the disease (1). Yet, it only characterizes transmission potential at the onset of the epidemic and varies geographically for a given infectious disease according to local healthcare provision, outbreak response, as well as socioeconomic and cultural factors. Furthermore, estimating R_0_ requires information about the natural history of the infectious disease. Thus, our ability to estimate reproduction numbers for novel infectious diseases is hindered by the paucity of information about their epidemiological characteristics and transmission mechanisms. More informative metrics could synthesize real-time information about the extent to which the epidemic is expanding over time. Such metrics would be particularly useful if they rely on minimal data on the outbreak’s trajectory (2).

#### Scope and definition

Our analysis is restricted to mainland China in this paper. A ‘province’ herein encompasses three different types of political sub-divisions of mainland China, namely, a province, a centrally (literally, ‘directly’) administered municipality (Beijing, Chongqing, Shanghai, and Tianjin) and an ‘ethnic minority’ autonomous region (Guangxi, Inner Mongolia, Ningxia, Tibet, and Xinjiang). Our analysis does not include the Hong Kong Special Administrative Region and the Macau Special Administrative Region, which are under the effective rule of the People’s Republic of China through the so-called ‘One Country, Two Systems’ political arrangements. Likewise, our analysis does not include Taiwan, which is *de facto* governed by a different government (the Republic of China).

#### Data sources

Daily cumulative incidence data were retrieved from provincial health commissions’ websites (Table S8). Data were double-checked against the cumulative national total published by the National Health Commission (3), data compiled by the Centre for Health Protection, Hong Kong, when available (4) and by John Hopkins University Center for Systems Science and Engineering (5). Whenever discrepancies arose, provincial government sources were deemed authoritative.

#### Doubling time calculation and its relationship with growth rate of an epidemic

As the epidemic grows, the times at which cumulative incidence doubles are given by 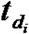 such that 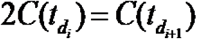 where 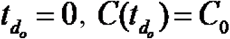 and *i =* 0,1,2,3, … *n*_*d*_ where ***n***_***d***_ is the total number of times cumulative incidence doubles. The actual sequence of “doubling times” are defined as follows:

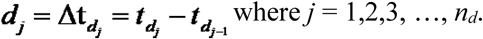

To quantify parameter uncertainty, we used parametric bootstrapping with a Poisson error structure around the harmonic mean of doubling times ***d***_***j***_ to obtain the 95% confidence interval. See references (6-8) for further details.

If we assume homogeneous mixing (equal probability of acquiring infection through contacts) and exponential growth, then, C(t_2_) = C(t_1_)exp(rt), and therefore, ln(C(t_2_)/C(t_1_)) = rt. When C(t_2_)/C(t_1_) = 2 and thus t is the doubling time, i.e. t = t_d_, ln2 = rt_d_. Therefore, the doubling time, t_d_, equals to (ln2)/r. See Vynnycky and White (9), panel 4.1, p.74 for further explanation.

#### Additional details on methods

Doubling time calculation was conducted using MATLAB R2019b (Mathworks, Natick, MA). Figures were created either using R version 3.6.2 (R Core Team) or MATLAB 2019b. Significance level in this manuscript was a priori decided to be α = 0.05.

### Additional information on our results and discussion

#### Cumulative incidence over time

From Figure S7 to Figure S10, we provided plots of cumulative incidence over time (left panel) and semi-log plots with log_10_-transformed cumulative incidence over time (right panel) for a total of 8 provinces with a relatively high number of cases, namely, the epicenter Hubei, followed by (in alphabetical order) Fujian, Guangdong, Heilongjiang, Henan, Hubei, Hunan, Jiangxi and Shandong. If the epidemic is growing exponentially, the log_10_-transformed cumulative incidence over time will be a linear curve. If social distancing would have an impact, the slope of the semi-log plot would decrease, indicating a decreasing epidemic growth rate.

#### Harmonic mean of the harmonic mean

In this study, we also presented the harmonic mean of the harmonic means of the estimates of the epidemic doubling times. The harmonic means of the epidemic doubling times are shorter than their arithmetic means. From January 20 through February 9, the harmonic mean of the harmonic means of the doubling times estimated ranged from 0.5 (95% CI, 0.2, 1.3) days for Guangxi, to 2.3 (95% CI, 2.3, 2.4) days for Hubei. The harmonic mean of the harmonic means of doubling times in mainland China except Hubei were 1.2 (95% CI, 1.0, 1.4) days.

#### Further discussion

The slowing-down of the epidemic as represented in increasing epidemic doubling times in our study is also consistent with a study by Benjamin F. Maier and Dirk Brockmann, “Effective containment explains sub-exponential growth in confirmed cases of recent COVID-19 outbreak in Mainland China” (pre-print available at arXiv. 2020:2002.07572). They also identified sub-exponential growth of the outbreak across provinces, as mass quarantine and restriction of travels across mainland China began since January 23, 2020.

#### Sensitivity analysis #1

We performed a sensitivity analysis by expanding our data analysis to the data since December 31, 2019, when Hubei first reported a cluster of pneumonia cases with unexplained etiology that turned out to be COVID-19. The only difference between the sensitivity analysis and the main analysis is the inclusion of Hubei and Guangdong data from December 31, 2019, through January 19, 2020, because nationwide reporting started on January 20, 2020. The only differences in results were found for Hubei and Guangdong. For Hubei, the harmonic mean of the arithmetic mean of the doubling times was 4.06 (95% CI, 3.85-4.33); the harmonic mean of the harmonic means of the doubling times for Hubei was 2.28 (95% CI, 2.08-2.56); and the cumulative incidence in Hubei doubled nine times from December 31, 2019, through February 9, 2020 (Table S5, Figures S3, S4, S12, S13, S14). The first doubling time of Hubei (Figure S3) was high, reflecting that real-time data was unavailable before mid-January. It was only by January 17, 2020, onwards when data reporting become increasingly transparent and timely.

#### Sensitivity analysis #2

We also performed a sensitivity analysis by restricting our data analysis to the data from January 23, 2020 through February 9, 2020, to allow for the time that all the other provinces to ramp up their testing. January 23 was also the day when the Chinese authorities to put the city of Wuhan on ‘lockdown’ and major inter-provincial travel restrictions were put in place. When we changed the start date of our study period from January 20 (main analysis) to January 23, 2020 (sensitivity analysis #2), the epidemic doubling time of the aggregate cumulative incidence of mainland China (except Hubei) increased from 1.79 (95% CI, 1.52, 2.25) to 2.90 (95% CI, 2.62, 3.24) (harmonic mean of the arithmetic means), and from 1.18 (95% CI, 0.96, 1.42) to 1.98 (95% CI, 1.82, 2.17) (harmonic mean of the harmonic means) (Table S7, Figure S5, S6). Apart from the epidemic doubling time of the aggregate cumulative incidence of mainland China (except Hubei), we did not observe significant differences by province between results in the main analysis and sensitivity analysis #2. Therefore, our results should be robust for the purpose of this study.

**Table S1.** Confirmed cases of COVID-19 (December 31, 2019 to January 19, 2020) by province in mainland China extracted from official government sources used for the sensitivity analysis.

|  | Dec | January |  |  |  |  |  |  |  |  |  |  |  |  |  |  |  |  |  |  |
| --- | --- | --- | --- | --- | --- | --- | --- | --- | --- | --- | --- | --- | --- | --- | --- | --- | --- | --- | --- | --- |
| **Locations<sup>1</sup>** | **31** | **1** | **2** | **3** | **4** | **5** | **6** | **7** | **8** | **9** | **10** | **11** | **12** | **13** | **14** | **15** | **16** | **17** | **18** | **19** |
| Mainland China (Excluding Hubei) (Sum of provincial reports) | NR | NR | NR | NR | NR | NR | NR | NR | NR | NR | NR | NR | NR | NR | NR | NR | NR | NR | NR | 1 |
| Mainland China (Including Hubei) (Sum of provincial reports) | 27 | NR | NR | 44 | NR | 59 | NR | NR | NR | NR | 41 | 41 | 41 | 41 | 41 | 41 | 45 | 62 | 121 | 199 |
| Mainland China (Including Hubei) (Sum by NCH)<sup>2</sup> | NA | NA | NA | NA | NA | NA | NA | NA | NA | NA | NA | NA | NA | NA | NA | NA | NA | NA | NA | NA |
| Hubei | 27 | NR | NR | 44 | NR | 59 | NR | NR | NR | NR | 41 | 41 | 41 | 41 | 41 | 41 | 45 | 62 | 121 | 198 |
| Guangdong | NR | NR | NR | NR | NR | NR | NR | NR | NR | NR | NR | NR | NR | NR | NR | NR | NR | NR | NR | 1 |

*Note.* NA, not applicable. NR, not reported. <sup>1</sup>Observations were collected directly from government official sites from each province in mainland China. If a press release included data reported at midnight and early morning, they were considered to belong to the previous day the data was reported. <sup>2</sup>Official national tally of cumulative case count of confirmed cases was first published by the National Health Commission of China on January 21, 2020 for January 20, 2020 (3).

**Table S2.** Confirmed cases of COVID-19 (January 20 to 31, 2020) by province in mainland China extracted from official government sources used for the main analysis and sensitivity analysis.

| Locations<sup>1</sup> | January |  |  |  |  |  |  |  |  |  |  |  |
| --- | --- | --- | --- | --- | --- | --- | --- | --- | --- | --- | --- | --- |
|  | 20 | 21 | 22 | 23 | 24 | 25 | 26 | 27 | 28 | 29 | 30 | 31 |
| Mainland China (Excluding Hubei) (Sum of provincial reports) | 26 | 71 | 145 | 291 | 585 | 923 | 1321 | 1802 | 2386 | 3126 | 3885 | 4637 |
| Mainland China (Including Hubei) (Sum of provincial reports) | 296 | 446 | 589 | 840 | 1314 | 1975 | 2744 | 4516 | 5940 | 7712 | 9691 | 11790 |
| Mainland China (Including Hubei) (Sum by NCH)<sup>2</sup> | 291 | 440 | 571 | 830 | 1287 | 1975 | 2744 | 4515 | 5974 | 7711 | 9692 | 11791 |
| Hubei | 270 | 375 | 444 | 549 | 729 | 1052 | 1423 | 2714 | 3554 | 4586 | 5806 | 7153 |
| Anhui | 0 | 1 | 9 | 15 | 39 | 60 | 70 | 106 | 152 | 200 | 237 | 297 |
| Beijing | 5 | 10 | 14 | 26 | 36 | 49 | 68 | 80 | 91 | 111 | 132 | 156 |
| Chongqing | 0 | 5 | 9 | 27 | 57 | 75 | 110 | 132 | 147 | 165 | 206 | 238 |
| Fujian | 0 | 0 | 1 | 5 | 10 | 18 | 35 | 59 | 82 | 101 | 120 | 144 |
| Gansu | 0 | 0 | 0 | 2 | 4 | 7 | 14 | 19 | 24 | 26 | 29 | 35 |
| Guangdong | 14 | 26 | 32 | 53 | 78 | 98 | 146 | 188 | 241 | 311 | 393 | 520 |
| Guangxi | 0 | 0 | 2 | 13 | 23 | 33 | 46 | 51 | 58 | 78 | 87 | 100 |
| Guizhou | 0 | 0 | 0 | 3 | 5 | 5 | 7 | 9 | 9 | 12 | 15 | 29 |
| Hainan | 0 | 0 | 4 | 8 | 11 | 20 | 27 | 33 | 40 | 46 | 49 | 57 |
| Hebei | 0 | 0 | 1 | 2 | 8 | 13 | 18 | 33 | 48 | 65 | 82 | 96 |
| Heilongjiang | 0 | 0 | 1 | 4 | 9 | 15 | 21 | 30 | 37 | 43 | 59 | 80 |
| Henan | 0 | 1 | 5 | 9 | 32 | 83 | 128 | 168 | 206 | 278 | 352 | 422 |
| Hunan | 0 | 1 | 4 | 9 | 43 | 69 | 100 | 143 | 221 | 277 | 332 | 389 |
| Inner Mongolia | 0 | 0 | 0 | 1 | 2 | 7 | 11 | 13 | 16 | 18 | 20 | 23 |
| Jiangsu | 0 | 0 | 1 | 9 | 18 | 31 | 47 | 70 | 99 | 129 | 168 | 202 |
| Jiangxi | 0 | 2 | 3 | 7 | 18 | 36 | 48 | 72 | 109 | 162 | 240 | 286 |
| Jilin | 0 | 0 | 1 | 3 | 4 | 4 | 6 | 8 | 9 | 14 | 14 | 17 |
| Liaoning | 0 | 0 | 2 | 4 | 12 | 19 | 22 | 30 | 36 | 41 | 45 | 60 |
| Ningxia | 0 | 0 | 1 | 2 | 3 | 4 | 7 | 11 | 12 | 17 | 21 | 26 |
| Qinghai | 0 | 0 | 0 | 0 | 0 | 1 | 4 | 6 | 6 | 6 | 8 | 9 |
| Shaanxi | 0 | 0 | 0 | 0 | 7 | 15 | 22 | 46 | 56 | 63 | 87 | 101 |
| Shandong | 0 | 1 | 1 | 1 | 21 | 39 | 63 | 87 | 87 | 145 | 178 | 202 |
| Shanghai | 2 | 9 | 16 | 20 | 33 | 40 | 53 | 66 | 80 | 101 | 128 | 153 |
| Shanxi | 0 | 0 | 1 | 1 | 6 | 9 | 13 | 20 | 27 | 35 | 39 | 47 |
| Sichuan | 0 | 2 | 5 | 15 | 28 | 44 | 69 | 90 | 108 | 142 | 177 | 207 |
| Tianjin | 0 | 2 | 4 | 5 | 8 | 10 | 14 | 23 | 25 | 27 | 32 | 32 |
| Tibet | 0 | 0 | 0 | 0 | 0 | 0 | 0 | 0 | 0 | 1 | 1 | 1 |
| Xinjiang | 0 | 0 | 0 | 2 | 3 | 4 | 5 | 10 | 13 | 14 | 17 | 18 |
| Yunnan | 0 | 1 | 1 | 2 | 5 | 11 | 19 | 26 | 51 | 70 | 80 | 91 |
| Zhejiang | 5 | 10 | 27 | 43 | 62 | 104 | 128 | 173 | 296 | 428 | 537 | 599 |

*Note.* <sup>1</sup>Observations were collected directly from government official sites from each province in mainland China. If a press release included data reported at midnight and early morning, they were considered to belong to the previous day the data was reported. <sup>2</sup>Data was collected of press releases of the National Health Commission of China (3).

**Table S3.** Confirmed cases of COVID-19 (February 1 to 9, 2020) by province in mainland China extracted from official government sources used for the main analysis and sensitivity analysis.

|  | February |  |  |  |  |  |  |  |  |
| --- | --- | --- | --- | --- | --- | --- | --- | --- | --- |
|  | 1 | 2 | 3 | 4 | 5 | 6 | 7 | 8 | 9 |
| **Locations<sup>1</sup>** |  |  |  |  |  |  |  |  |  |
| Mainland China (Excluding Hubei) (Sum of provincial reports) | 5396 | 6031 | 6910 | 7646 | 8352 | 9049 | 9614 | 10098 | 10507 |
| Mainland China (Including Hubei) (Sum of provincial reports) | 14381 | 17208 | 20432 | 24324 | 28017 | 31161 | 34567 | 37198 | 40138 |
| Mainland China (Including Hubei) (Sum by NCH)<sup>2</sup> | 14380 | 17205 | 20438 | 24324 | 28018 | 31161 | 34546 | 37198 | 40171 |
| Hubei | 9074 | 11177 | 13522 | 16678 | 19665 | 22112 | 24953 | 27100 | 29631 |
| Anhui | 340 | 408 | 480 | 530 | 591 | 665 | 733 | 779 | 830 |
| Beijing | 183 | 212 | 228 | 253 | 274 | 297 | 315 | 326 | 337 |
| Chongqing | 262 | 300 | 337 | 366 | 389 | 411 | 426 | 446 | 468 |
| Fujian | 159 | 179 | 194 | 205 | 215 | 224 | 239 | 250 | 261 |
| Gansu | 40 | 51 | 55 | 57 | 62 | 67 | 71 | 79 | 83 |
| Guangdong | 604 | 683 | 797 | 870 | 944 | 1018 | 1075 | 1120 | 1131 |
| Guangxi | 111 | 127 | 139 | 150 | 168 | 172 | 183 | 195 | 210 |
| Guizhou | 38 | 46 | 56 | 64 | 69 | 77 | 89 | 96 | 99 |
| Hainan | 63 | 70 | 79 | 89 | 100 | 111 | 122 | 128 | 136 |
| Hebei | 104 | 113 | 126 | 135 | 157 | 171 | 195 | 206 | 218 |
| Heilongjiang | 95 | 118 | 155 | 190 | 227 | 277 | 295 | 307 | 331 |
| Henan | 493 | 566 | 675 | 764 | 851 | 914 | 981 | 1033 | 1073 |
| Hunan | 463 | 521 | 593 | 661 | 711 | 772 | 803 | 838 | 879 |
| Inner Mongolia | 27 | 34 | 35 | 42 | 46 | 50 | 52 | 54 | 58 |
| Jiangsu | 236 | 271 | 308 | 341 | 373 | 408 | 439 | 468 | 492 |
| Jiangxi | 333 | 391 | 476 | 548 | 600 | 661 | 698 | 740 | 771 |
| Jilin | 23 | 31 | 42 | 54 | 59 | 65 | 69 | 78 | 80 |
| Liaoning | 64 | 73 | 74 | 81 | 89 | 94 | 99 | 105 | 108 |
| Ningxia | 28 | 31 | 34 | 34 | 40 | 43 | 45 | 45 | 49 |
| Qinghai | 9 | 13 | 15 | 17 | 18 | 18 | 18 | 18 | 18 |
| Shaanxi | 116 | 128 | 142 | 165 | 173 | 184 | 195 | 208 | 213 |
| Shandong | 225 | 246 | 270 | 298 | 343 | 379 | 407 | 435 | 466 |
| Shanghai | 177 | 193 | 208 | 233 | 254 | 269 | 281 | 292 | 295 |
| Shanxi | 56 | 66 | 74 | 81 | 90 | 96 | 104 | 115 | 119 |
| Sichuan | 231 | 254 | 282 | 301 | 321 | 344 | 363 | 386 | 405 |
| Tianjin | 45 | 48 | 60 | 67 | 69 | 81 | 88 | 90 | 94 |
| Tibet | 1 | 1 | 1 | 1 | 1 | 1 | 1 | 1 | 1 |
| Xinjiang | 21 | 24 | 29 | 32 | 36 | 39 | 42 | 45 | 49 |
| Yunnan | 99 | 109 | 117 | 122 | 128 | 135 | 138 | 140 | 141 |
| Zhejiang | 661 | 724 | 829 | 895 | 954 | 1006 | 1048 | 1075 | 1092 |

*Note.* <sup>1</sup>Observations were collected directly from government official sites from each province in mainland China. If a press release included data reported at midnight and early morning, they were considered to belong to the previous day the data was reported. <sup>2</sup>Data was collected of press releases of the National Health Commission of China (3).

**Table S4.** Main analysis: Doubling times of COVID-19 cumulative incidence and their harmonic mean of the arithmetic means of the doubling times and harmonic mean of the harmonic means of the doubling times (95% Confidence interval) by province in mainland China from January 20 through February 9, 2020.

|  |  | Mainland China<br>(Except Hubei) | Hubei | Anhui | Beijing | Chongqing | Fujian | Gansu | Guangdong | Guangxi | Guizhou | Hainan |
| --- | --- | --- | --- | --- | --- | --- | --- | --- | --- | --- | --- | --- |
| Harmonic mean of arithmetic means |  | 1.79<br>(1.52-2.25) | 2.54<br>(2.44-2.64) | 2.56<br>(2.16-3.11) | 2.49<br>(1.89-3.38) | 2.22<br>(1.53-3.22) | 1.71<br>(1.15-2.52) | 2.56<br>(2.00-3.78) | 2.47<br>(1.97-3.20) | 1.92<br>(1.45-3.09) | 2.71<br>(1.90-3.90) | 2.91<br>(1.91-3.89) |
| Harmonic mean of harmonic means |  | 1.18<br>(0.96-1.42) | 2.34<br>(2.27-2.41) | 1.72<br>(1.13-2.67) | 1.48<br>(0.63-2.70) | 1.23<br>(0.67-1.96) | 0.82<br>(0.46-1.41) | 1.36<br>(0.76-2.86) | 2.01<br>(1.53-2.54) | 0.48<br>(0.22-1.34) | 1.88<br>(0.81-3.28) | 1.52<br>(0.65-2.99) |
| Times doubled | 1 | 0.59 | 2.91 | 2.12 | 1.00 | 2.05 | 0.25 | 1 | 1.33 | 0.18 | 2.5 | 1.00 |
|  | 2 | 0.86 | 2.16 | 0.75 | 1.5 | 0.56 | 0.5 | 1.14 | 1.79 | 0.36 | 3.5 | 1.55 |
|  | 3 | 0.98 | 1.5 | 2.18 | 1.8 | 0.82 | 0.85 | 1.26 | 2.17 | 0.76 | 1.64 | 2.28 |
|  | 4 | 1 | 2.17 | 1.77 | 2.7 | 1.71 | 1.15 | 4.1 | 2.38 | 1.6 | 2.56 | 5.31 |
|  | 5 | 1.3 | 3.03 | 4.03 | 4.14 | 3.58 | 1.07 | 5.9 | 2.76 | 3.4 | 5.8 | 6.86 |
|  | 6 | 1.98 | 3.43 | 4.9 | 7.31 | 4.82 | 1.39 |  | 4.92 | 4.78 |  |  |
|  | 7 | 2.55 |  |  |  |  | 3.12 |  |  |  |  |  |
|  | 8 | 4.53 |  |  |  |  | 9.21 |  |  |  |  |  |
|  |  | Hebei | Heilongjiang | Henan | Hunan | Inner Mongolia | Jiangsu | Jiangxi | Jilin | Liaoning | Ningxia | Qinghai |
| Harmonic mean of arithmetic means |  | 1.88<br>(1.57-2.72) | 1.93<br>(1.76-2.36) | 1.81<br>(1.35-2.05) | 1.42<br>(1.24-2.04) | 2.37<br>(1.80-3.67) | 2.43<br>(1.77-3.26) | 1.68<br>(1.45-2.33) | 2.64<br>(2.13-3.50) | 2.10<br>(1.45-3.30) | 2.54<br>(1.76-4.33) | 2.50<br>(1.50-5.00) |
| Harmonic mean doubling time |  | 1.04<br>(0.67-1.93) | 1.08<br>(0.62-1.98) | 0.81<br>(0.56-1.12) | 0.71<br>(0.47-1.13) | 1.17<br>(0.67-2.67) | 1.93<br>(1.35-2.63) | 1.13<br>(0.71-1.71) | 1.48<br>(0.66-3.03) | 1.05<br>(0.54-1.94) | 1.59<br>(0.73-3.07) | 1.00<br>(0.38-3.87) |
| Times doubled | 1 | 1 | 0.33 | 0.25 | 0.33 | 1 | 2 | 1.25 | 0.5 | 1 | 1 | 0.33 |
|  | 2 | 0.33 | 0.67 | 0.5 | 0.67 | 0.4 | 1.31 | 0.84 | 2.5 | 0.5 | 2 | 0.67 |
|  | 3 | 0.67 | 0.8 | 1 | 0.8 | 0.85 | 1.75 | 0.72 | 2 | 1.07 | 1.25 | 4 |
|  | 4 | 1.6 | 1.36 | 0.55 | 0.4 | 2.75 | 2.32 | 0.96 | 3.66 | 2.76 | 2.55 | 4.5 |
|  | 5 | 1.33 | 2.12 | 0.7 | 0.47 | 4.71 | 4.07 | 1.89 | 2.43 | 4.67 | 4.53 |  |
|  | 6 | 2.01 | 2.95 | 0.62 | 1.13 |  |  | 1.69 | 3.74 |  |  |  |
|  | 7 | 5.28 | 3.04 | 1.38 | 1.85 |  |  | 1.99 |  |  |  |  |
|  | 8 |  | 3.31 | 2.69 | 1.97 |  |  | 4.16 |  |  |  |  |
|  | 9 |  |  | 3.57 | 4.22 |  |  |  |  |  |  |  |
|  | 10 |  |  | 6.56 |  |  |  |  |  |  |  |  |
|  |  | Shaanxi | Shandong | Shanghai | Shanxi | Sichuan | Tianjin | Tibet | Xinjiang | Yunnan | Zhejiang |  |
| Harmonic mean of the arithmetic means |  | 2.82<br>(2.12-9.97) | 1.68<br>(1.42-2.39) | 2.19<br>(1.88-2.68) | 2.31<br>(1.67-3.25) | 1.83<br>(1.39-2.70) | 2.78<br>(2.07-4.06) | Not applied | 3.05<br>(2.06-4.75) | 2.05<br>(1.34-2.72) | 1.91<br>(1.60-2.51) |  |
| Harmonic mean doubling time |  | 2.04<br>(1.28-3.01) | 0.48<br>(0.28-1.15) | 0.77<br>(0.34-1.73) | 1.22<br>(0.68-2.51) | 0.96<br>(0.51-1.75) | 1.69<br>(0.80-3.55) | Not applied | 1.91<br>(0.83-4.46) | 1.25<br>(0.89-1.81) | 1.20<br>(0.74-1.70) |  |
| Times doubled | 1 | 1.33 | 2.05 | 0.28 | 1.2 | 0.66 | 1 |  | 2 | 2 | 1 |  |
|  | 2 | 2.24 | 0.1 | 0.57 | 0.4 | 0.64 | 2 |  | 1.6 | 0.66 | 0.58 |  |
|  | 3 | 3.76 | 0.19 | 1.15 | 1.06 | 0.77 | 2.22 |  | 3.06 | 0.84 | 1.23 |  |
|  | 4 |  | 0.41 | 1.92 | 1.76 | 1.18 | 4.78 |  | 5.34 | 1.12 | 1.61 |  |
|  | 5 |  | 0.86 | 2.92 | 2.2 | 1.55 | 3.57 |  |  | 1.61 | 2.29 |  |
|  | 6 |  | 1.43 | 3.16 | 4.18 | 2.78 |  |  |  | 1.45 | 1.47 |  |
|  | 7 |  | 2.66 | 6.13 |  | 4.49 |  |  |  | 7.32 | 3.48 |  |
|  | 8 |  | 4.71 |  |  |  |  |  |  |  |  |  |

**Table S5.** Sensitivity analysis #1 (1 of 2 tables): Doubling times of COVID-19 cumulative incidence and their harmonic mean of the arithmetic means of the doubling times and harmonic mean of the harmonic means of the doubling times (95% Confidence interval) by province in mainland China from December 31, 2019 through February 9, 2020: Mainland China (Except Hubei), Hubei, and from Anhui to Qinghai.

|  |  | Mainland China<br>(Except Hubei) | Hubei | Anhui | Beijing | Chongqing | Fujian | Gansu | Guangdong | Guangxi | Guizhou | Hainan |
| --- | --- | --- | --- | --- | --- | --- | --- | --- | --- | --- | --- | --- |
| Harmonic mean of arithmetic means |  | 1.34<br>(1.28-1.52) | 4.06<br>(3.85-4.33) | 2.57<br>(2.12-3.00) | 2.51<br>(1.99-3.26) | 2.22<br>(1.60-3.23) | 1.82<br>(1.18-2.55) | 2.55<br>(1.83-3.79) | 1.88<br>(1.74-2.19) | 1.93<br>(1.47-2.96) | 2.78<br>(2.00-3.97) | 2.92<br>(1.97-4.25) |
| Harmonic mean of harmonic means |  | 0.29<br>(0.15-0.59) | 2.28<br>(2.08-2.56) | 1.76<br>(1.21-2.40) | 1.60<br>(0.93-2.70) | 1.23<br>(0.74-1.88) | 0.83<br>(0.47-1.42) | 1.33<br>(0.70-2.62) | 0.44<br>(0.25-1.13) | 0.49<br>(0.22-1.29) | 1.98<br>(1.09-3.53) | 1.55<br>(0.60-3.29) |
| Times doubled | 1 | 0.04 | 17.33 | 2.12 | 1.00 | 2.05 | 0.25 | 1.00 | 0.07 | 0.18 | 2.50 | 1.00 |
|  | 2 | 0.08 | 1.22 | 0.75 | 1.5 | 0.56 | 0.5 | 1.14 | 0.16 | 0.36 | 3.5 | 1.55 |
|  | 3 | 0.15 | 2 | 2.18 | 1.8 | 0.82 | 0.85 | 1.26 | 0.3 | 0.76 | 1.64 | 2.28 |
|  | 4 | 0.33 | 3.04 | 1.77 | 2.7 | 1.71 | 1.15 | 4.1 | 0.63 | 1.6 | 2.56 | 5.31 |
|  | 5 | 0.53 | 2.11 | 4.03 | 4.14 | 3.58 | 1.07 | 5.9 | 1.84 | 3.4 | 5.8 | 6.86 |
|  | 6 | 0.73 | 1.23 | 4.9 | 7.31 | 4.82 | 1.39 |  | 1.44 | 4.78 |  |  |
|  | 7 | 0.92 | 2.61 |  |  |  | 3.12 |  | 2.18 |  |  |  |
|  | 8 | 0.98 | 3.13 |  |  |  | 9.21 |  | 2.59 |  |  |  |
|  | 9 | 1.01 | 3.87 |  |  |  |  |  | 2.72 |  |  |  |
|  | 10 | 1.48 |  |  |  |  |  |  | 6.17 |  |  |  |
|  | 11 | 2.17 |  |  |  |  |  |  |  |  |  |  |
|  | 12 | 2.88 |  |  |  |  |  |  |  |  |  |  |
|  | 13 | 5.55 |  |  |  |  |  |  |  |  |  |  |
|  |  | Hebei | Heilongjiang | Henan | Hunan | Inner Mongolia | Jiangsu | Jiangxi | Jilin | Liaoning | Ningxia | Qinghai |
| Harmonic mean of arithmetic means |  | 1.89<br>(1.55-2.74) | 1.96<br>(1.76-2.26) | 1.80<br>(1.31-2.10) | 1.41<br>(1.26-1.99) | 2.37<br>(1.82-3.57) | 2.45<br>(1.75-3.31) | 1.72<br>(1.44-2.36) | 2.67<br>(2.13-3.50) | 2.16<br>(1.49-3.53) | 2.58<br>(1.72-4.43) | 2.64<br>(1.79-5.00) |
| Harmonic mean of harmonic means |  | 1.07<br>(0.66-1.90) | 1.12<br>(0.66-1.97) | 0.77<br>(0.48-1.14) | 0.73<br>(0.48-1.15) | 1.15<br>(0.65-2.71) | 1.92<br>(1.31-2.68) | 1.17<br>(0.81-1.74) | 1.60<br>(0.70-3.11) | 1.06<br>(0.50-2.45) | 1.67<br>(0.87-3.65) | 0.96<br>(0.39-3.69) |
| Times doubled | 1 | 1.00 | 0.33 | 0.25 | 0.33 | 1.00 | 2.00 | 1.25 | 0.50 | 1.00 | 1.00 | 0.33 |
|  | 2 | 0.33 | 0.67 | 0.5 | 0.67 | 0.4 | 1.31 | 0.84 | 2.5 | 0.5 | 2 | 0.67 |
|  | 3 | 0.67 | 0.8 | 1 | 0.8 | 0.85 | 1.75 | 0.72 | 2 | 1.07 | 1.25 | 4 |
|  | 4 | 1.6 | 1.36 | 0.55 | 0.4 | 2.75 | 2.32 | 0.96 | 3.66 | 2.76 | 2.55 | 4.5 |
|  | 5 | 1.33 | 2.12 | 0.7 | 0.47 | 4.71 | 4.07 | 1.89 | 2.43 | 4.67 | 4.53 |  |
|  | 6 | 2.01 | 2.95 | 0.62 | 1.13 |  |  | 1.69 | 3.74 |  |  |  |
|  | 7 | 5.28 | 3.04 | 1.38 | 1.85 |  |  | 1.99 |  |  |  |  |
|  | 8 |  | 3.31 | 2.69 | 1.97 |  |  | 4.16 |  |  |  |  |
|  | 9 |  |  | 3.57 | 4.22 |  |  |  |  |  |  |  |
|  | 10 |  |  | 6.56 |  |  |  |  |  |  |  |  |

**Table S6.** Sensitivity analysis #1 (2 of 2 tables): Doubling times of COVID-19 cumulative incidence and their harmonic mean of the arithmetic means of the doubling times and harmonic mean of the harmonic means of the doubling times (95% Confidence interval) by province in mainland China from December 31, 2019 through February 9, 2020: from Shaanxi to Zhejiang.

|  |  | Shaanxi | Shandong | Shanghai | Shanxi | Sichuan | Tianjin | Tibet | Xinjiang | Yunnan | Zhejiang |
| --- | --- | --- | --- | --- | --- | --- | --- | --- | --- | --- | --- |
|  | Harmonic mean of arithmetic means | 2.77<br>(2.06-3.93) | 1.68<br>(1.41-2.36) | 2.21<br>(1.91-2.78) | 2.12<br>(1.67-3.00) | 1.79<br>(1.40-2.65) | 2.75<br>(2.10-3.89) | Not applied | 3.09<br>(2.12-4.89) | 2.10<br>(1.42-2.78) | 1.90<br>(1.59-2.55) |
|  | Harmonic mean of harmonic means | 2.03<br>(1.27-2.93) | 0.48<br>(0.30-1.11) | 0.82<br>(0.40-1.83) | 1.26<br>(0.68-2.60) | 0.96<br>(0.62-1.73) | 1.67<br>(0.78-3.38) | Not applied | 1.98<br>(0.80-4.69) | 1.28<br>(0.80-1.93) | 1.23<br>(0.77-1.72) |
| Times doubled | 1 | 1.33 | 2.05 | 0.28 | 1.20 | 0.66 | 1.00 |  | 2.00 | 2 | 1.00 |
|  | 2 | 2.24 | 0.1 | 0.57 | 0.4 | 0.64 | 2 |  | 1.6 | 0.66 | 0.58 |
|  | 3 | 3.76 | 0.19 | 1.15 | 1.06 | 0.77 | 2.22 |  | 3.06 | 0.84 | 1.23 |
|  | 4 |  | 0.41 | 1.92 | 1.76 | 1.18 | 4.78 |  | 5.34 | 1.12 | 1.61 |
|  | 5 |  | 0.86 | 2.92 | 2.2 | 1.55 | 3.57 |  |  | 1.61 | 2.29 |
|  | 6 |  | 1.43 | 3.16 | 4.18 | 2.78 |  |  |  | 1.45 | 1.47 |
|  | 7 |  | 2.66 | 6.13 |  | 4.49 |  |  |  | 7.32 | 3.48 |
|  | 8 |  | 4.71 |  |  |  |  |  |  |  |  |

**Table S7.** Sensitivity analysis #2: Doubling times of COVID-19 cumulative incidence and their harmonic mean of the arithmetic means of the doubling times and harmonic mean of the harmonic means of the doubling times (95% Confidence interval) by province in mainland China from January 23, 2020 through February 9, 2020.

|  |  | Mainland China<br>(Except Hubei) | Hubei | Anhui | Beijing | Chongqing | Fujian | Gansu | Guangdong | Guangxi | Guizhou | Hainan |
| --- | --- | --- | --- | --- | --- | --- | --- | --- | --- | --- | --- | --- |
| Harmonic mean of arithmetic means |  | 2.9<br>(2.62-3.24) | 2.46<br>(2.37-2.55) | 2.54<br>(2.12-2.99) | 3.46<br>(2.77-4.57) | 3.11<br>(2.38-4.17) | 2.03<br>(1.29-3.10) | 2.54<br>(1.80-3.89) | 2.91<br>(2.40-3.61) | 3.26<br>(2.37-4.22) | 2.67<br>(1.85-3.92) | 3.43<br>(2.57-4.62) |
| Harmonic mean of harmonic means |  | 1.98<br>(1.82-2.17) | 2.25<br>(2.18-2.33) | 1.47<br>(0.90-2.29) | 3.03<br>(2.23-3.99) | 1.87<br>(1.28-2.80) | 1.26<br>(0.69-2.01) | 1.27<br>(0.66-2.84) | 2.65<br>(2.14-3.10) | 2.23<br>(1.33-3.29) | 1.73<br>(0.67-3.40) | 2.31<br>(1.40-3.71) |
| Times doubled | 1 | 1.01 | 2.12 | 0.62 | 2.15 | 0.90 | 1.00 | 1.00 | 2.16 | 1.30 | 2.50 | 1.55 |
|  | 2 | 1.59 | 1.47 | 1.38 | 3.50 | 2.04 | 1.11 | 1.14 | 2.29 | 2.84 | 3.50 | 2.28 |
|  | 3 | 2.30 | 2.22 | 2.30 | 4.21 | 4.37 | 1.09 | 1.26 | 2.79 | 4.22 | 1.64 | 5.31 |
|  | 4 | 3.21 | 3.03 | 3.74 |  | 7.99 | 1.71 | 4.10 | 4.45 | 8.50 | 2.56 | 6.86 |
|  | 5 | 6.41 | 3.45 | 3.96 |  |  | 4.13 | 5.90 |  |  | 5.80 |  |
|  |  | Hebei | Heilongjiang | Henan | Hunan | Inner Mongolia | Jiangsu | Jiangxi | Jilin | Liaoning | Ningxia | Qinghai |
| Harmonic mean of arithmetic means |  | 1.91<br>(1.42-2.83) | 2.21<br>(1.74-2.81) | 1.87<br>(1.50-2.40) | 1.89<br>(1.48-2.77) | 2.39<br>(1.84-4.00) | 2.31<br>(1.80-3.10) | 1.89<br>(1.44-2.52) | 3.01<br>(2.14-4.06) | 2.44<br>(1.49-4.00) | 2.68<br>(1.70-4.50) | 3.21<br>(2.25-5.67) |
| Harmonic mean of harmonic means |  | 0.81<br>(0.39-1.78) | 1.47<br>(0.77-2.51) | 0.85<br>(0.48-1.37) | 0.75<br>(0.41-1.27) | 1.16<br>(0.66-3.02) | 1.60<br>(1.06-2.36) | 1.18<br>(0.62-1.86) | 2.44<br>(1.29-3.73) | 0.99<br>(0.37-2.46) | 1.88<br>(0.94-3.96) | 1.8<br>(0.80-5.08) |
| Times doubled | 1 | 0.33 | 0.80 | 0.39 | 0.26 | 1.00 | 1.00 | 0.63 | 3.00 | 0.50 | 2.00 | 2.33 |
|  | 2 | 0.67 | 1.36 | 0.68 | 0.53 | 0.40 | 1.31 | 0.92 | 2.59 | 1.07 | 1.25 | 0.67 |
|  | 3 | 1.60 | 2.12 | 0.71 | 1.30 | 0.85 | 1.75 | 1.78 | 3.53 | 2.76 | 2.55 | 4.00 |
|  | 4 | 1.33 | 2.95 | 1.62 | 1.92 | 2.75 | 2.32 | 1.72 | 2.38 | 4.67 | 4.53 | 4.50 |
|  | 5 | 2.01 | 3.04 | 2.73 | 2.19 | 4.71 | 4.07 | 1.74 |  |  |  |  |
|  | 6 | 5.28 | 3.31 | 3.96 | 4.56 |  |  | 3.88 |  |  |  |  |
|  |  | Shaanxi | Shandong | Shanghai | Shanxi | Sichuan | Tianjin | Tibet | Xinjiang | Yunnan | Zhejiang |  |
| Harmonic mean of arithmetic means |  | 3.44<br>(2.76-4.40) | 1.43<br>(1.18-2.14) | 3.08<br>(2.52-4.07) | 1.93<br>(1.50-3.09) | 2.61<br>(1.89-3.60) | 3.17<br>(2.16-4.59) | Not applied | 3.05<br>(2.10-4.67) | 1.82<br>(1.20-2.87) | 2.37<br>(1.87-3.14) |  |
| Harmonic mean of harmonic means |  | 2.84<br>(1.82-4.05) | 0.24<br>(0.14-0.60) | 2.61<br>(1.72-3.59) | 0.71<br>(0.32-1.88) | 1.82<br>(1.28-2.57) | 2.12<br>(0.79-4.34) | Not applied | 1.86<br>(0.83-4.40) | 1.03<br>(0.56-1.77) | 1.98<br>(1.73-2.41) |  |
| Times doubled | 1 | 3.33 | 0.05 | 2.00 | 0.20 | 1.12 | 2.00 |  | 2.00 | 0.66 | 1.57 |  |
|  | 2 | 2.24 | 0.10 | 3.00 | 0.40 | 1.51 | 1.66 |  | 1.60 | 0.84 | 2.4 |  |
|  | 3 | 3.76 | 0.20 | 3.29 | 1.06 | 2.72 | 4.95 |  | 3.06 | 1.12 | 1.39 |  |
|  | 4 |  | 0.40 |  | 1.76 | 4.04 | 5.30 |  | 5.34 | 1.61 | 4.06 |  |
|  | 5 |  | 0.86 |  | 2.20 |  |  |  |  | 1.45 |  |  |
|  | 6 |  | 1.43 |  | 4.18 |  |  |  |  | 7.32 |  |  |
|  | 7 |  | 2.66 |  |  |  |  |  |  |  |  |  |
|  | 8 |  | 4.71 |  |  |  |  |  |  |  |  |  |

**Table S8.**
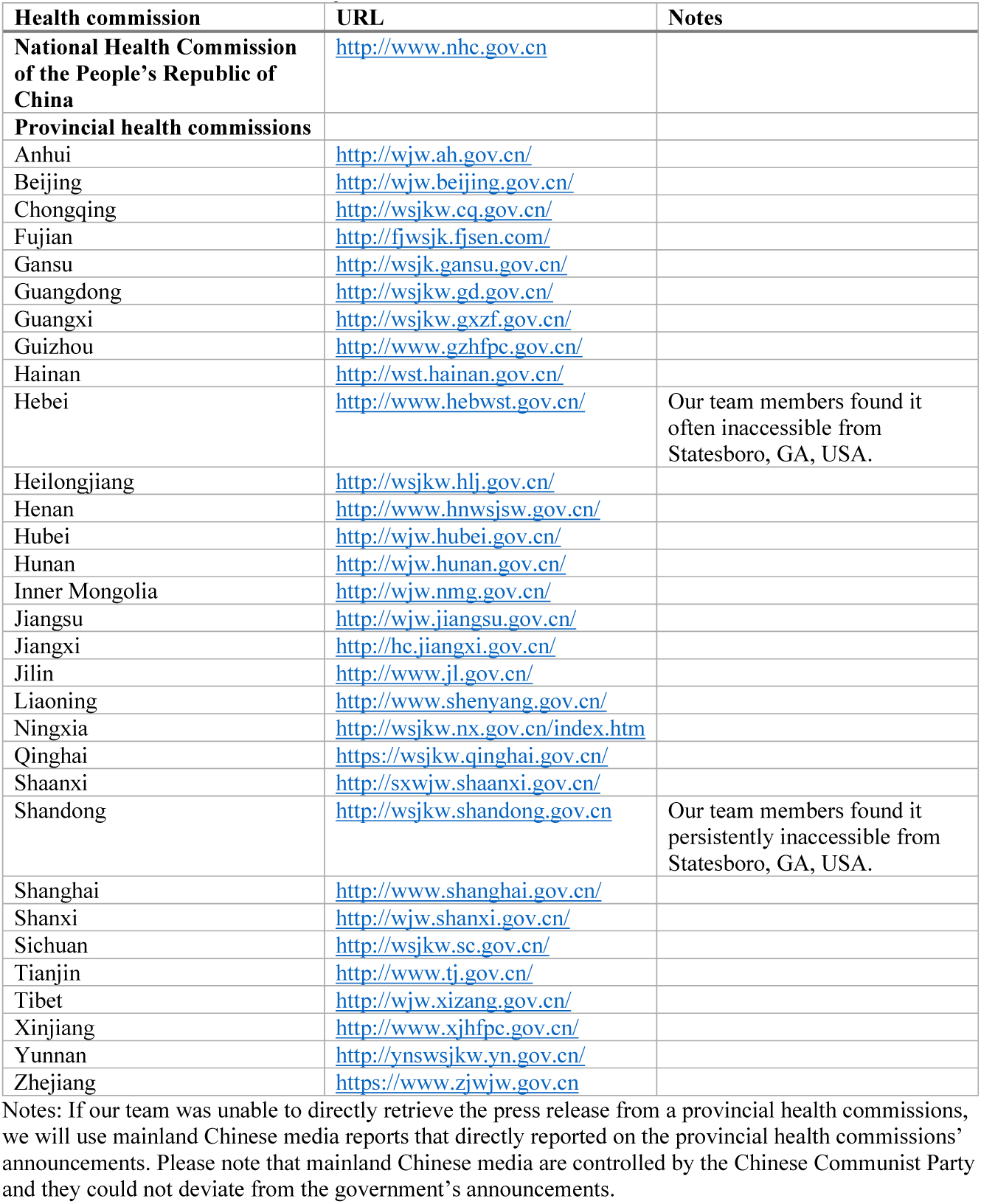
Websites of national and provincial health commissions in mainland China

**Figure S1.**
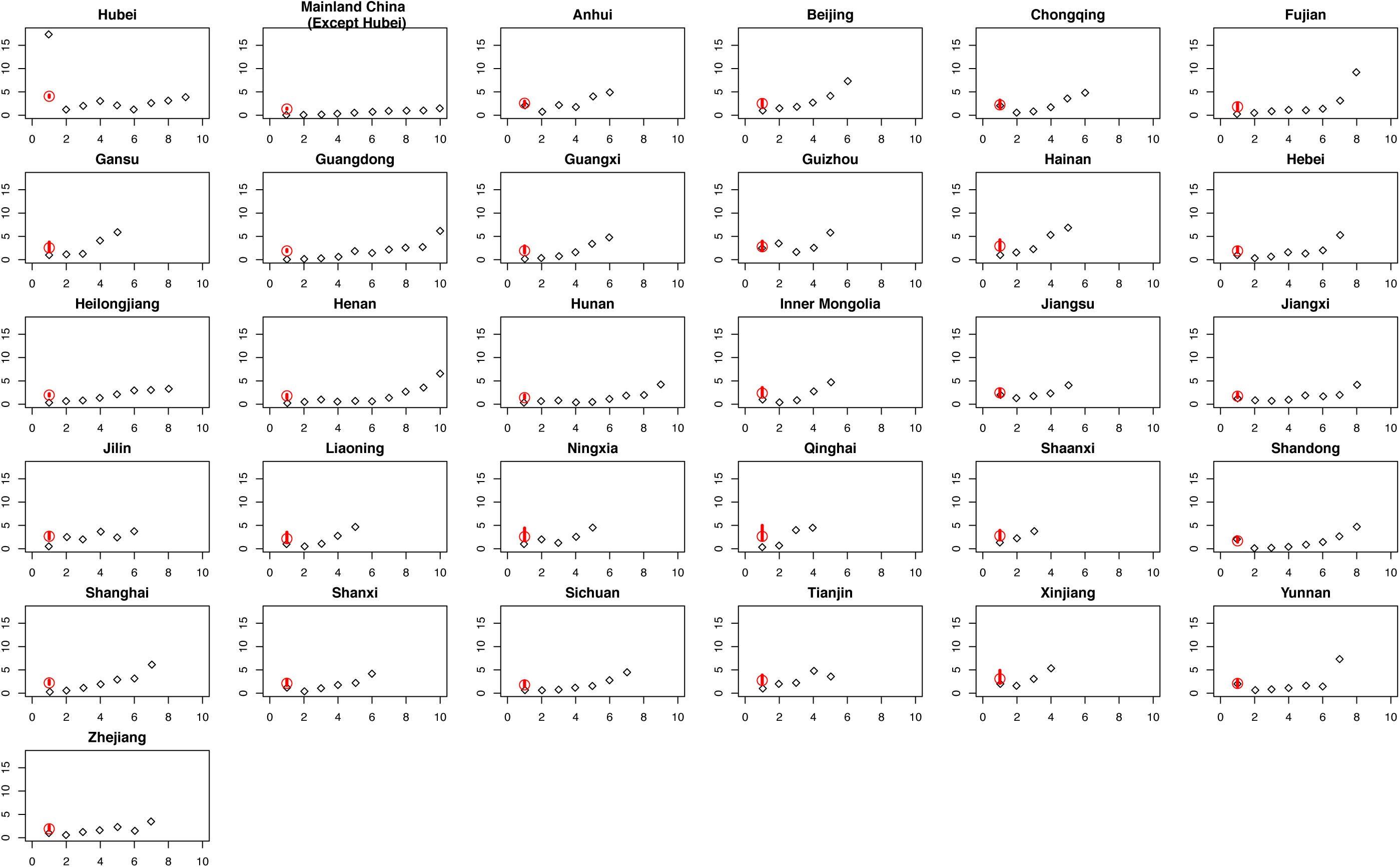
Main analysis: The harmonic mean of the arithmetic means of COVID-19 epidemic doubling times (red circle) with 95% confidence interval (red bar) of the doubling times (days), and their values (black diamond) by the number of times the reported cumulative incidence doubles by province within mainland China from January 20, 2020 through February 9, 2020. Each panel represents a province except the panel representing “Mainland China (except Hubei)” that is the aggregate of all other provinces in mainland China, except Hubei. Doubling time for Tibet is not available, because there had only been 1 confirmed case in Tibet as of February 9, 2020.

**Figure S2.**
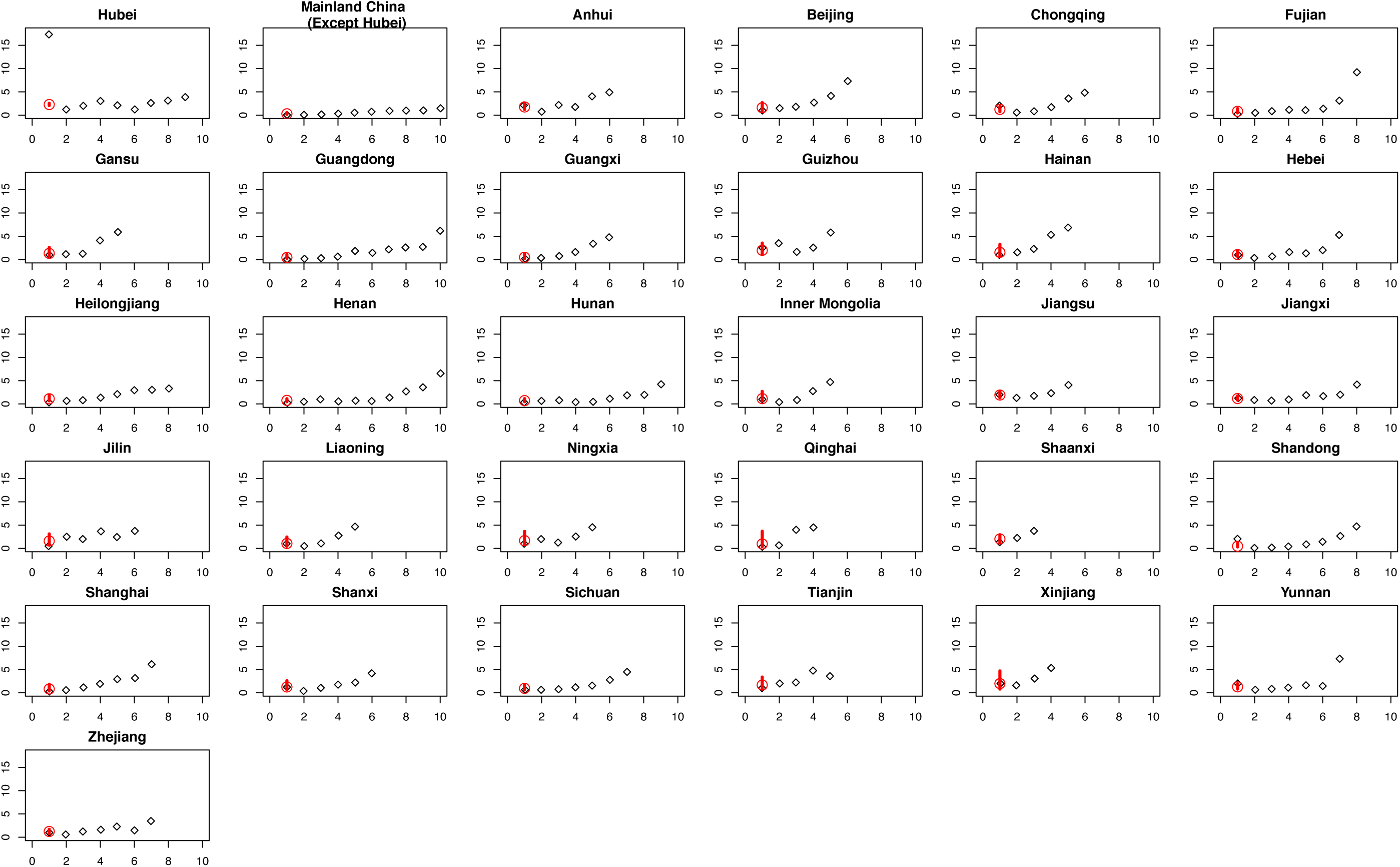
Main analysis: The harmonic mean of the harmonic means of COVID-19 epidemic doubling times (red circle) with 95% confidence interval (red bar) of the doubling times (days), and their values (black diamond) by the number of times the reported cumulative incidence doubles by province within mainland China from January 20, 2020 through February 9, 2020. Each panel represents a province except the panel representing “Mainland China (except Hubei)” that is the aggregate of all other provinces in mainland China, except Hubei. Doubling time for Tibet is not available, because there had only been 1 confirmed case in Tibet as of February 9, 2020.

**Figure S3.**
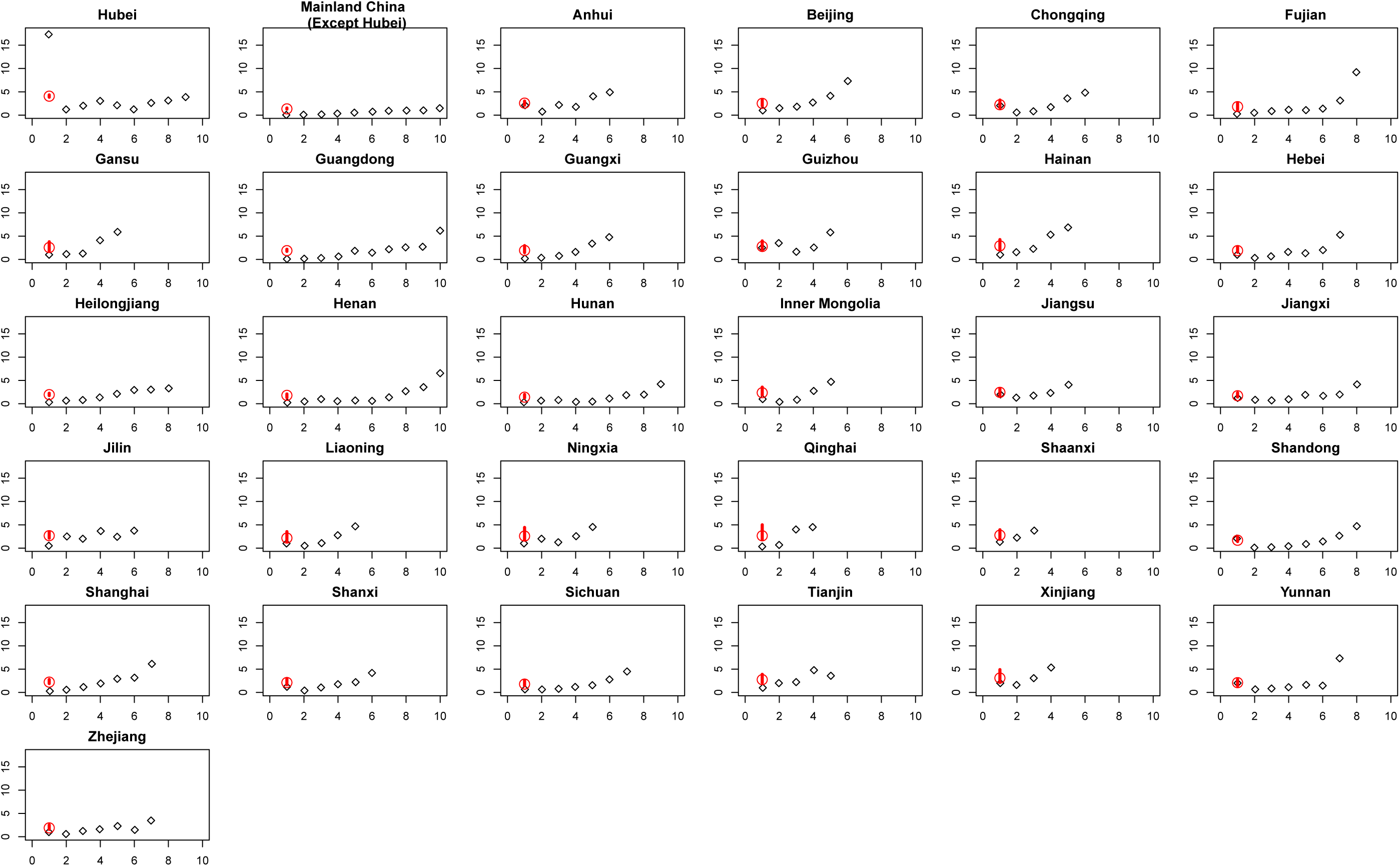
Sensitivity analysis #1: The harmonic mean of the arithmetic means of COVID-19 doubling times (red circle) with 95% confidence interval (red bar) of the doubling times (days), and their values (black diamond) by the number of times the reported cumulative incidence doubles by province within mainland China from December 31, 2019 through February 9, 2020. Each panel represents a province except the panel representing “Mainland China (except Hubei)” that is the aggregate of all other provinces in mainland China, except Hubei. Doubling time for Tibet is not available, because there had only been 1 confirmed case in Tibet as of February 9, 2020.

**Figure S4.**
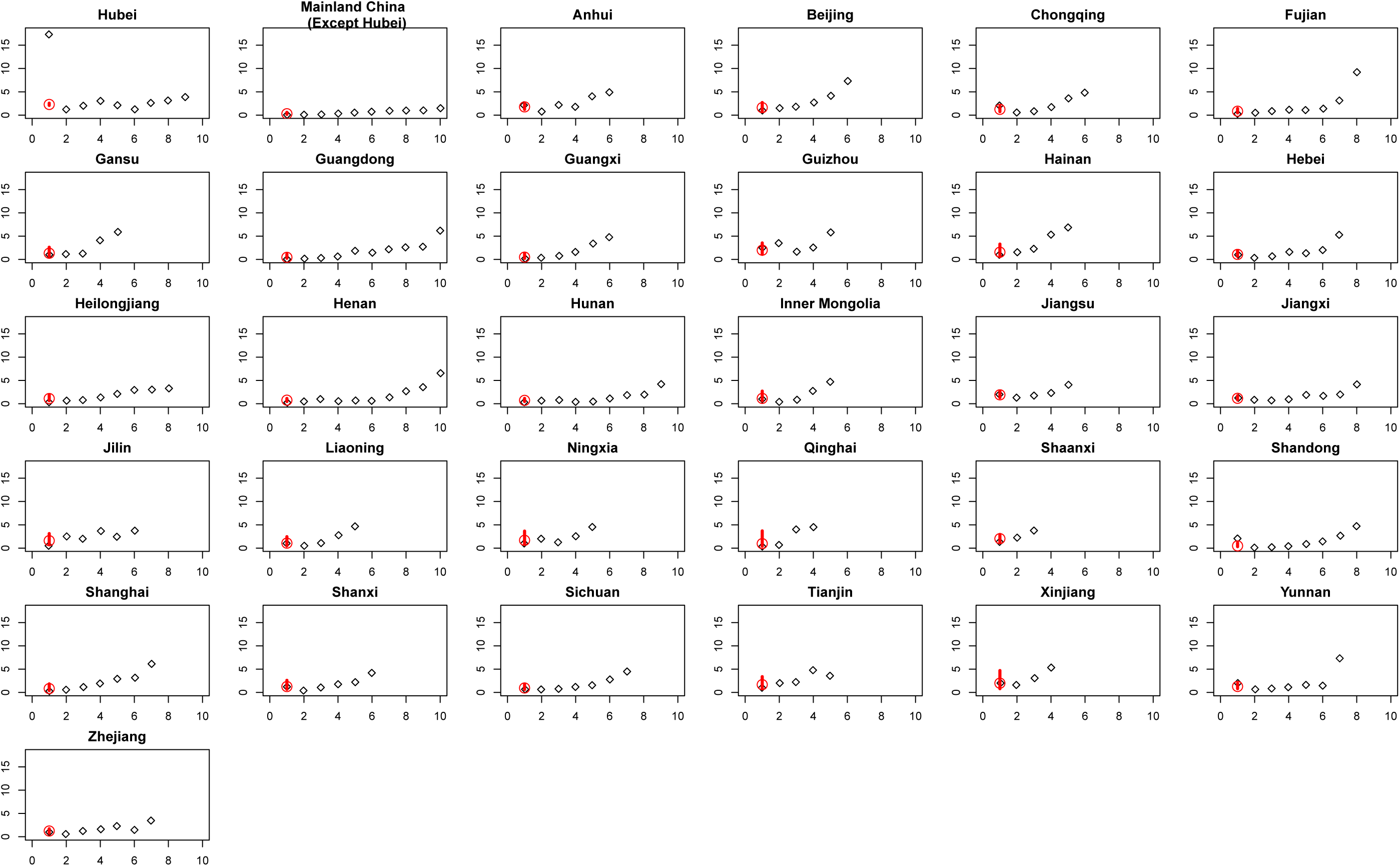
Sensitivity analysis #1: The harmonic mean of the harmonic means of COVID-19 doubling times (red circle) with 95% confidence interval (red bar) of the doubling times (days), and their values (black diamond) by the number of times the reported cumulative incidence doubles by province within mainland China from December 31, 2019 through February 9, 2020. Each panel represents a province except the panel representing “Mainland China (except Hubei)” that is the aggregate of all other provinces in mainland China, except Hubei. Doubling time for Tibet is not available, because there had only been 1 confirmed case in Tibet as of February 9, 2020.

**Figure S5.**
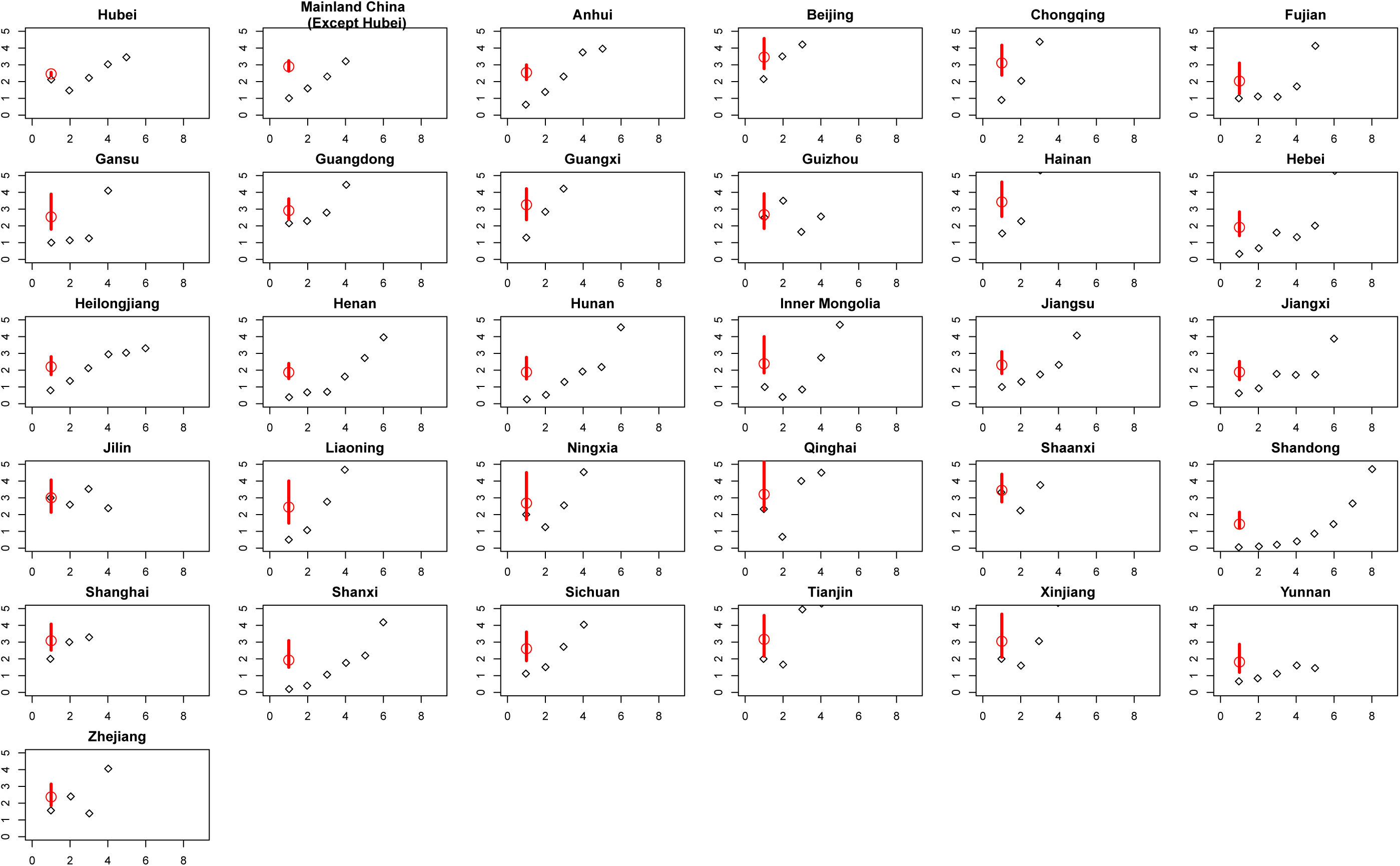
Sensitivity analysis #2: The harmonic mean of the arithmetic means of COVID-19 doubling times (red circle) with 95% confidence interval (red bar) of the doubling times (days), and their values (black diamond) by the number of times the reported cumulative incidence doubles by province within mainland China from January 23, 2020 through February 9, 2020. Each panel represents a province except the panel representing “Mainland China (except Hubei)” that is the aggregate of all other provinces in mainland China, except Hubei. Doubling time for Tibet is not available, because there had only been 1 confirmed case in Tibet as of February 9, 2020.

**Figure S6.**
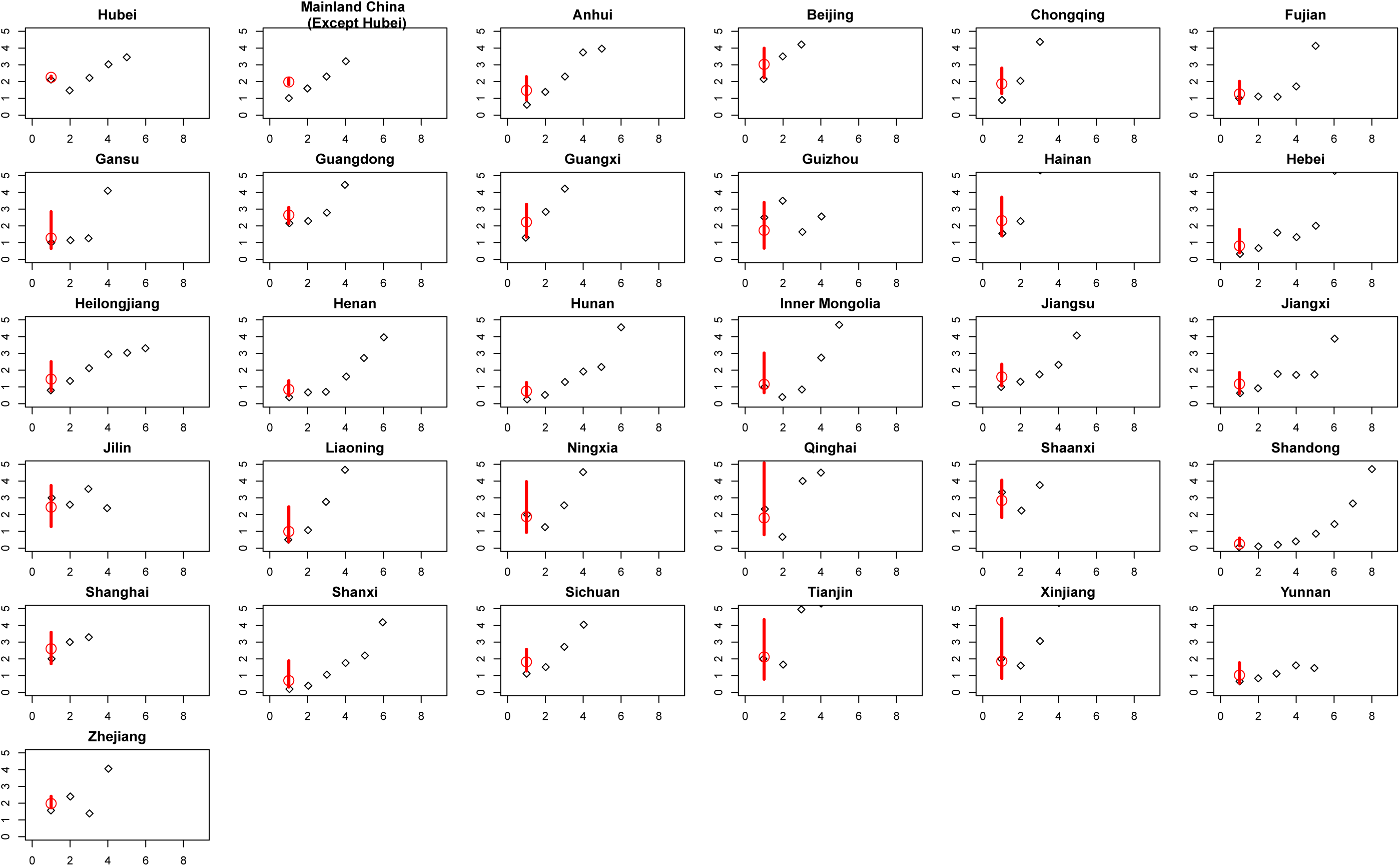
Sensitivity analysis #2: The harmonic mean of the harmonic means of COVID-19 doubling times (red circle) with 95% confidence interval (red bar) of the doubling times (days), and their values (black diamond) by the number of times the reported cumulative incidence doubles by province within mainland China from January 23, 2020 through February 9, 2020. Each panel represents a province except the panel representing “Mainland China (except Hubei)” that is the aggregate of all other provinces in mainland China, except Hubei. Doubling time for Tibet is not available, because there had only been 1 confirmed case in Tibet as of February 9, 2020.

**Figure S7.**
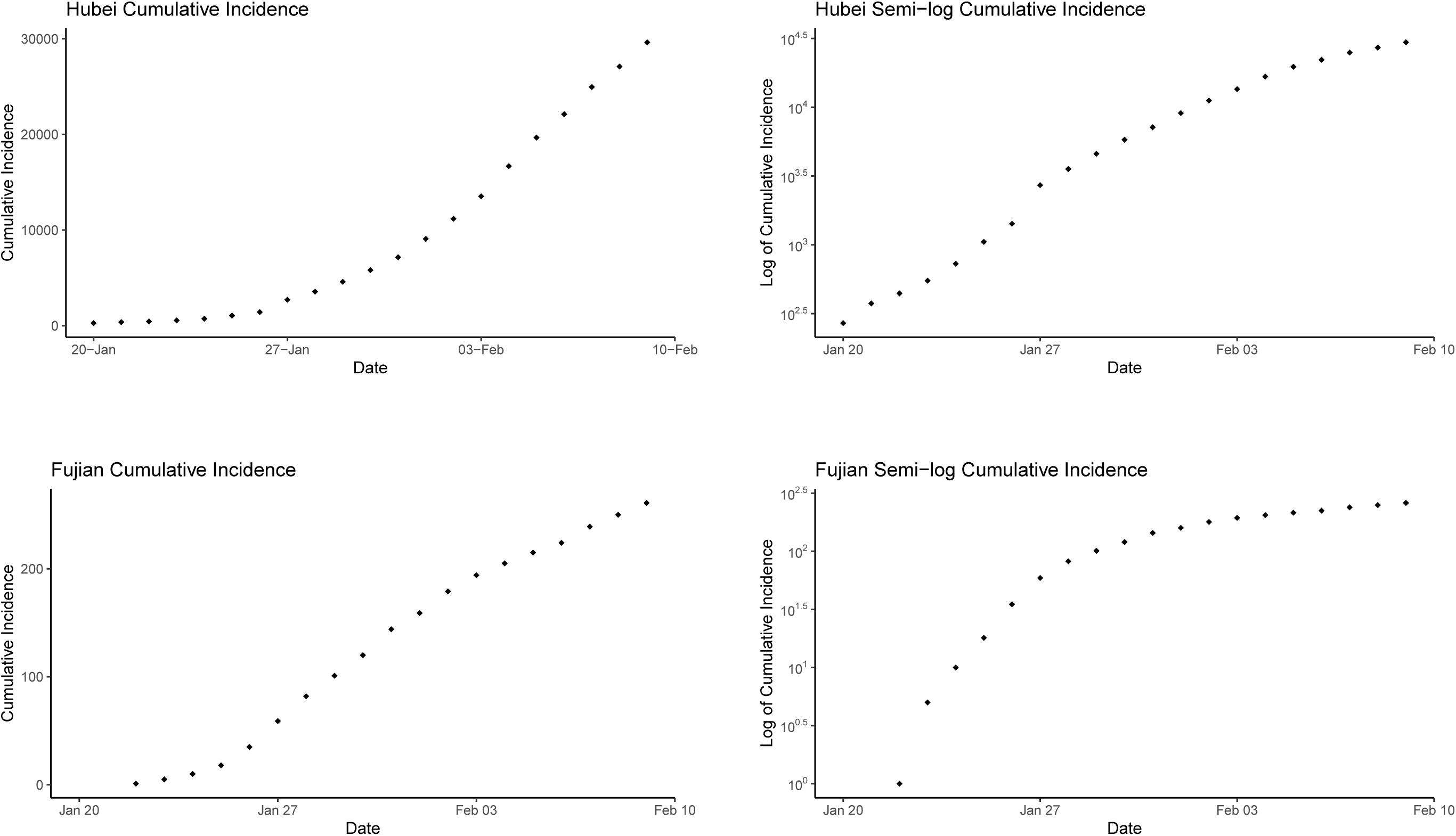
Cumulative incidence and log_10_ cumulative incidence over time (date) for Hubei (upper panel) and Fujian (lower panel).

**Figure S8.**
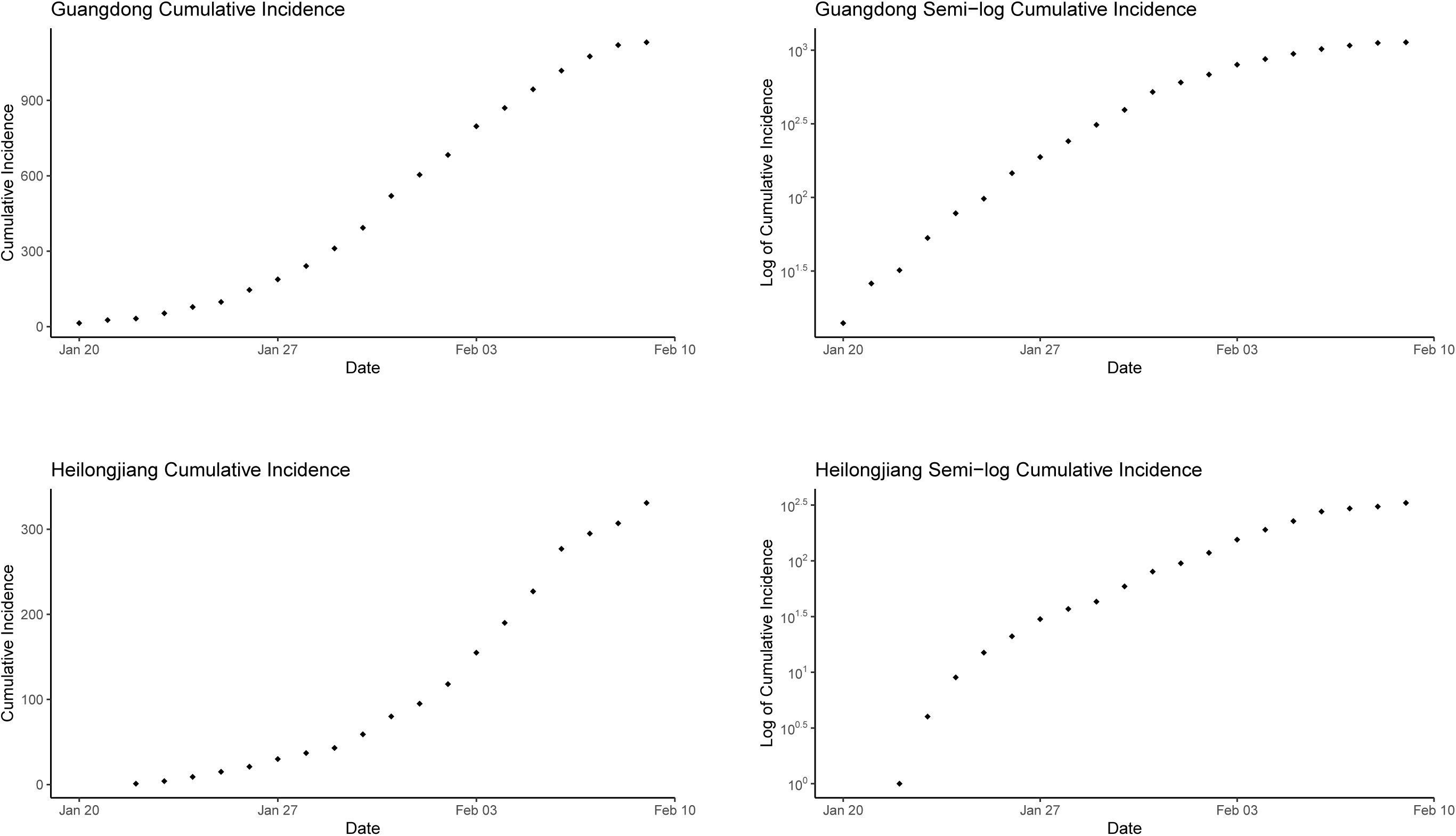
Cumulative incidence and log_10_ cumulative incidence over time (date) for Guangdong (upper panel) and Heilongjiang (lower panel).

**Figure S9.**
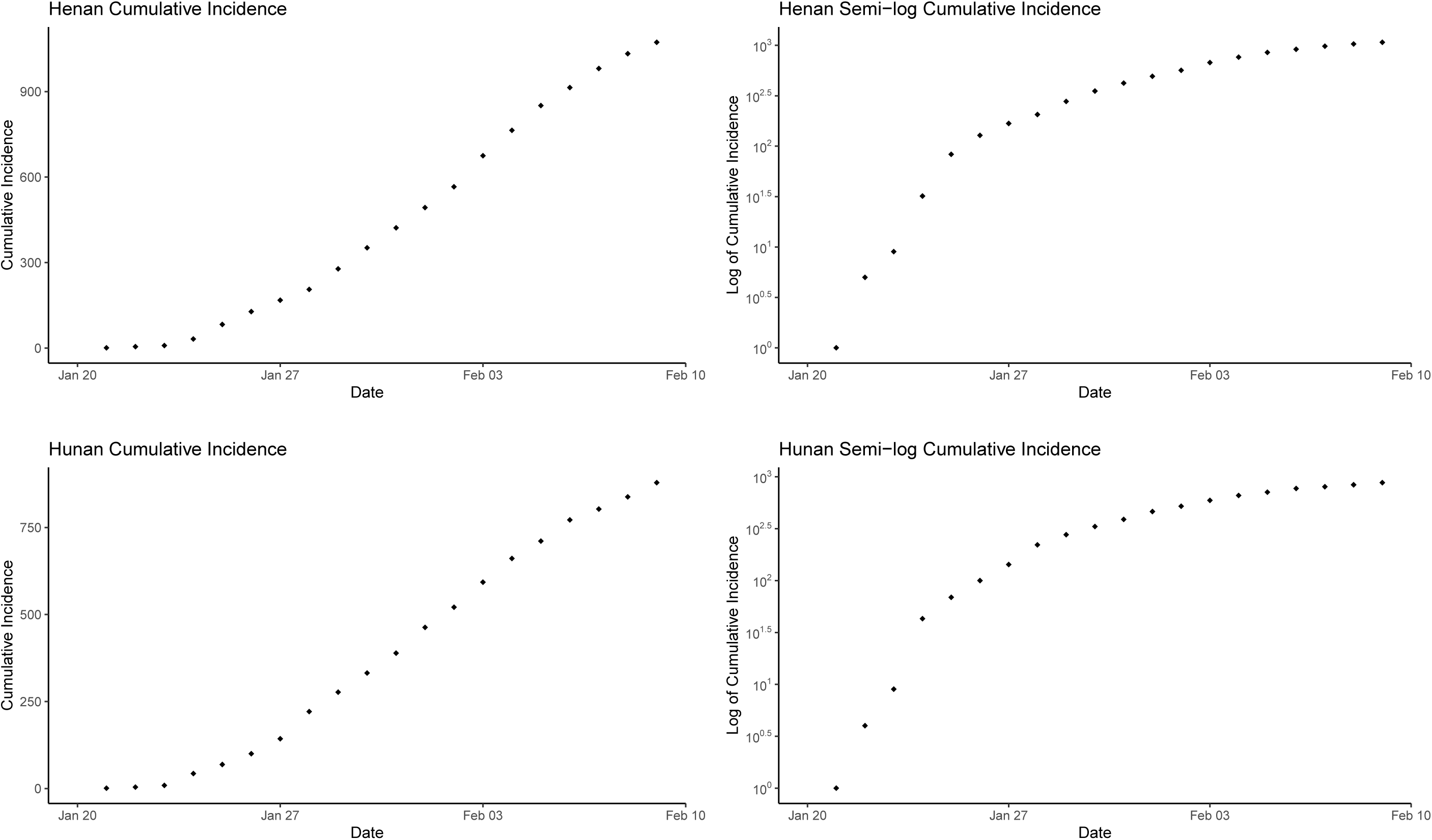
Cumulative incidence and log_10_ cumulative incidence over time (date) for Henan (upper panel) and Hunan (lower panel).

**Figure S10.**
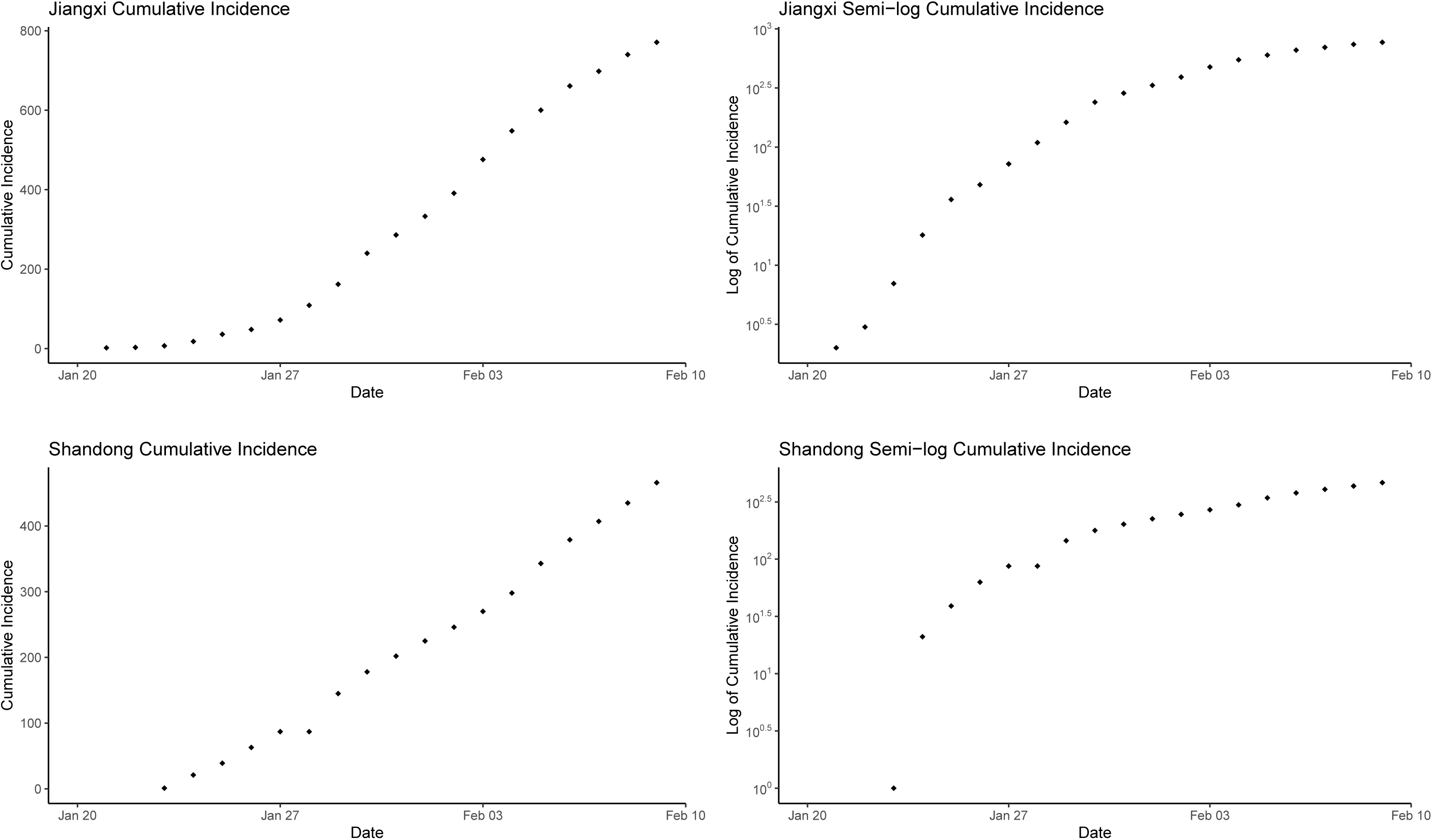
Cumulative incidence and log_10_ cumulative incidence over time (date) for Jiangxi (upper panel) and Shandong (lower panel).

**Figure S11.**
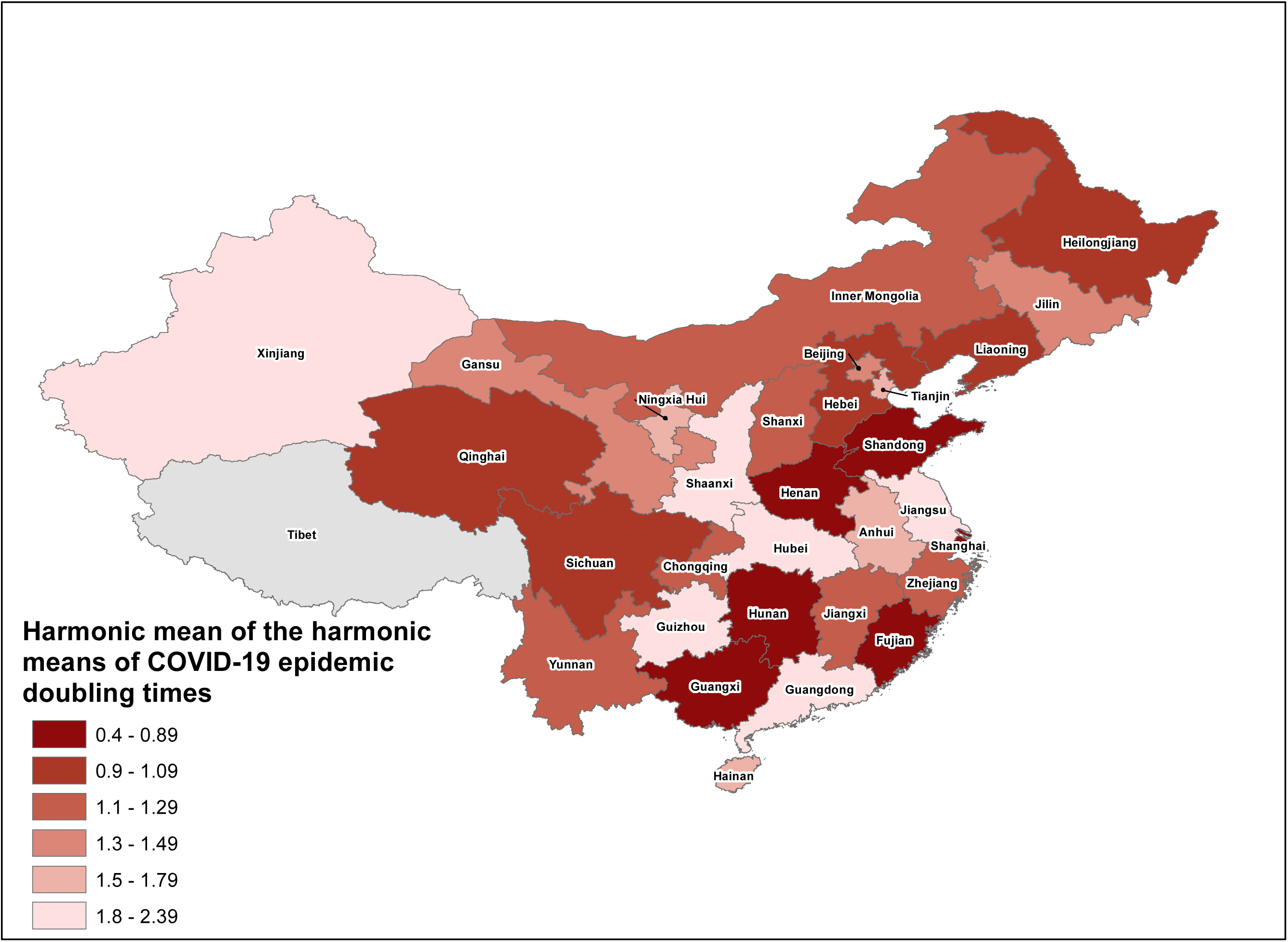
Main analysis: The map of the harmonic mean of the harmonic means of COVID-19 by province in mainland China, from January 20, 2020 through February 9, 2020.

**Figure S12.**
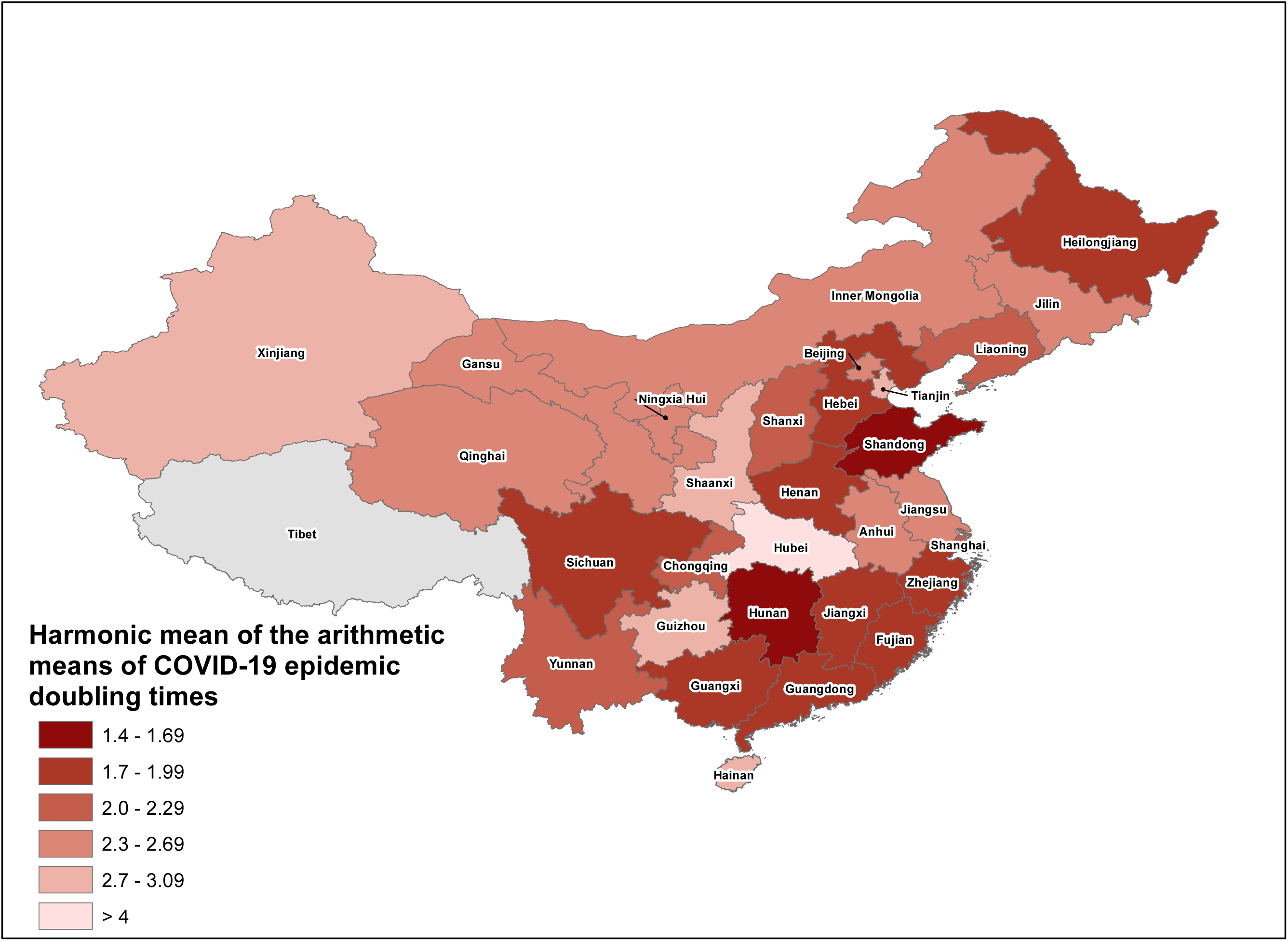
Sensitivity analysis #1: The map of the harmonic mean of the arithmetic means of COVID-19 by province in mainland China, from December 31, 2019 through February 9, 2020.

**Figure S13.**
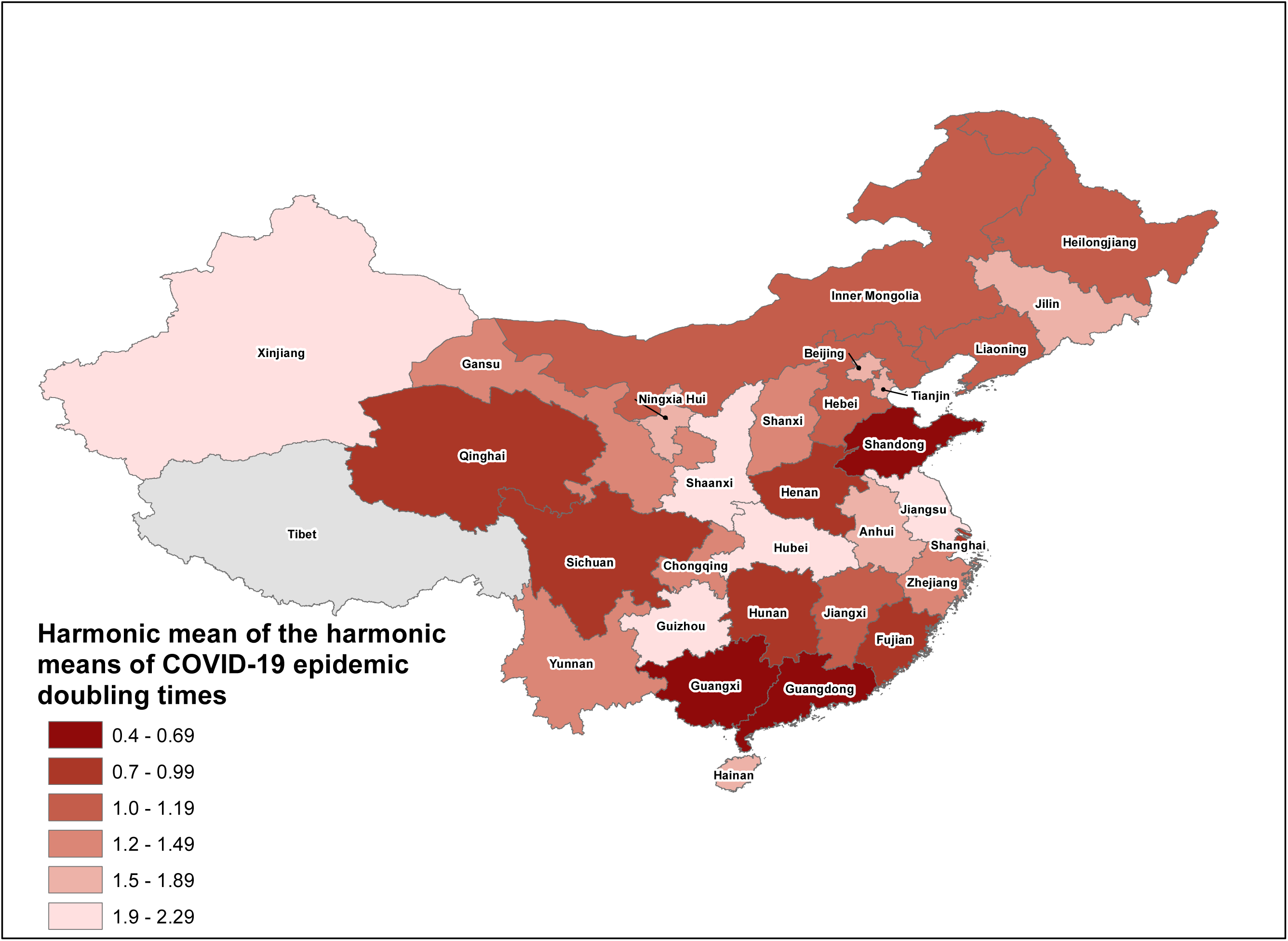
Sensitivity Analysis #1: The map of the harmonic mean of the harmonic means of COVID-19 by province in mainland China, from December 31, 2019 through February 9, 2020.

**Figure S14.**
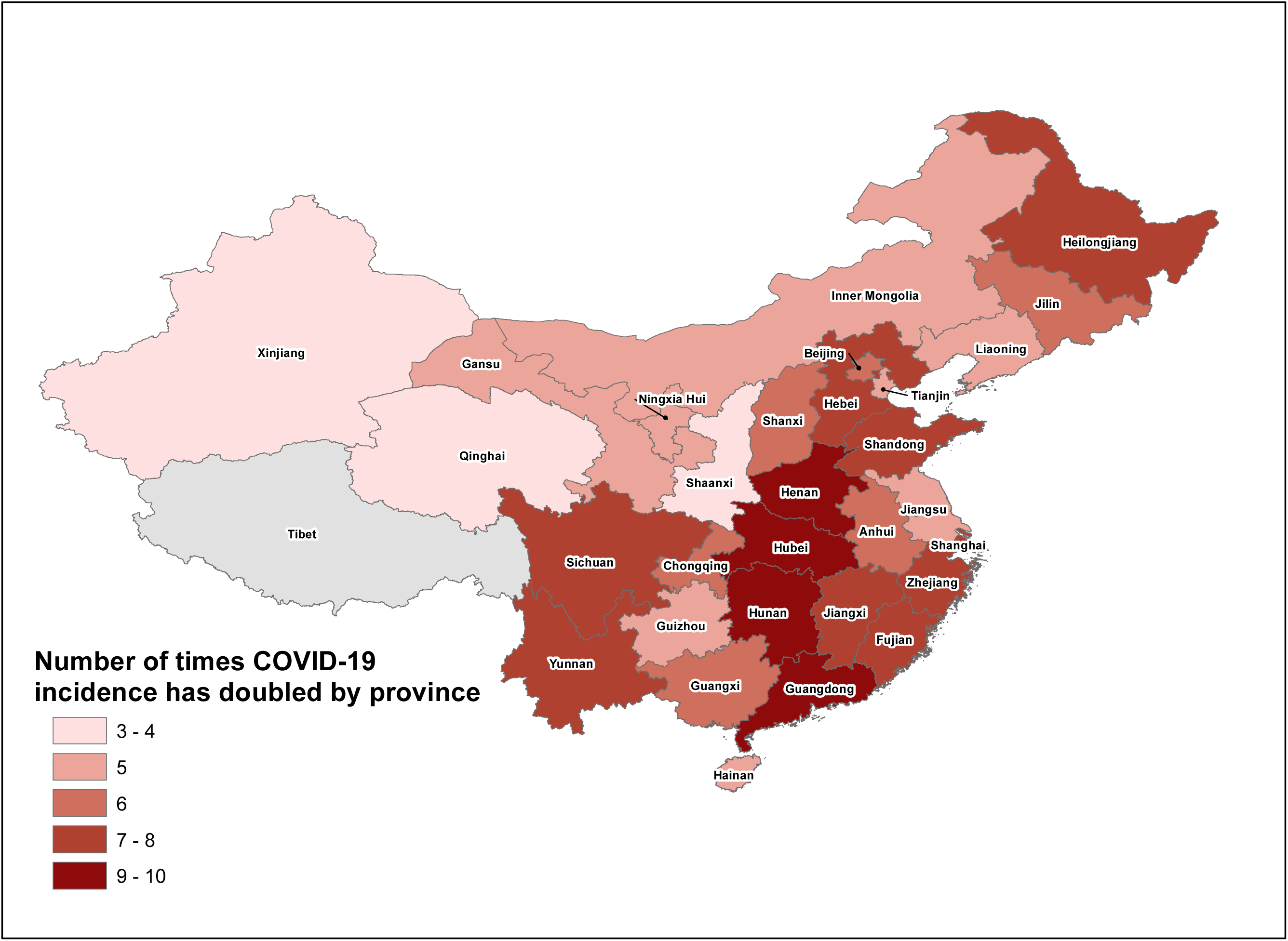
Sensitivity analysis #1: The map of the number of times the COVID-19 outbreak has doubled by province in mainland China, from December 31, 2019 through February 9, 2020.

## Authors’ contributions

Project management: Dr. Gerardo Chowell, Dr. Isaac Chun-Hai Fung and Ms. Kamalich Muniz-Rodriguez

Manuscript writing: Dr. Isaac Chun-Hai Fung and Dr. Gerardo Chowell

Manuscript editing and data interpretation: Ms. Kamalich Muniz-Rodriguez, Dr. Gerardo Chowell, Dr. Isaac Chun-Hai Fung, Dr. Lone Simonsen, and Dr. Cecile Viboud

MATLAB code and methods of doubling time estimation: Dr. Gerardo Chowell

Doubling time calculation using MATLAB and presentation of results: Ms. Kamalich Muniz-Rodriguez, Dr. Gerardo Chowell and Dr. Isaac Chun-Hai Fung

Statistical analysis in R: Dr. Isaac Chun-Hai Fung, Ms. Kamalich Muniz-Rodriguez

Data management and quality check of epidemic data entry: Ms. Kamalich Muniz-Rodriguez, Dr. Isaac Chun-Hai Fung

Curation of epidemic data for countries and territories outside mainland China (including Hong Kong, Macao and Taiwan): Ms. Kamalich Muniz-Rodriguez and Ms. Sylvia K. Ofori

Curation of epidemic data for provinces in mainland China: Ms. Manyun Liu (from the early reports, up to Jan 24, 2020 data), Ms. Po-Ying Lai (since Jan 25, 2020 data to today), Mr. Chi-Hin Cheung (since Jan 27, 2020 data to today), and Ms. Kamalich Muniz-Rodriguez and Dr. Isaac Chun-Hai Fung (whenever there is a back-log).

Retrieval of epidemic data from official websites (downloading and archiving of China’s national and provincial authorities’ press releases): Ms. Manyun Liu and Dr. Dongyu Jia (at the very beginning of our project)

Retrieval of statistical data from the official website of National Bureau of Statistics of the People’s Republic of China: Mr. Chi-Hin Cheung

Retrieval of publicly available statistical data from various sources: Ms. Yiseul Lee, Dr. Isaac Chun-Hai Fung

Table S8: Ms. Manyun Liu, Dr. Isaac Chun-Hai Fung Map creation: Ms. Kimberlyn M. Roosa

Assistance provided to Dr. Fung and Ms. Muniz-Rodriguez: Ms. Sylvia K. Ofori

## References

1. Anderson RM, May RM. Infectious diseases of humans. Oxford: Oxford University Press; 1991.

2. Drake JM, Bakach I, Just MR, O’Regan SM, Gambhir M, Fung IC-H. Transmission Models of Historical Ebola Outbreaks. Emerging Infectious Disease journal. 2015;21(8):1447.

3. Vynnycky E, White RG. An Introduction to Infectious Disease Modelling. Oxford: Oxford University Press; 2010.

4. Muniz-Rodriguez K, Fung IC-H, Ferdosi SR, Ofori SK, Lee Y, Tariq A, et al. Transmission potential of SARS-CoV-2 in Iran, 2020. Emerging Infectious Disease journal. 2020;26(8):DOI: https://doi.org/10.3201/eid2608.200536.

5. Li Q, Guan X, Wu P, Wang X, Zhou L, Tong Y, et al. Early Transmission Dynamics in Wuhan, China, of Novel Coronavirus-Infected Pneumonia. N Engl J Med. 2020 Mar 26 382(13):1199–207.

6. Du Z, Wang L, Cauchemez S, Xu X, Wang X, Cowling BJ, et al. Risk for Transportation of Coronavirus Disease from Wuhan to Other Cities in China. Emerging Infectious Disease journal. 2020 May 17;26(5):1049–52.

7. Wu JT, Leung K, Leung GM. Nowcasting and forecasting the potential domestic and international spread of the 2019-nCoV outbreak originating in Wuhan, China: a modelling study. Lancet. 2020 Feb 29;395(10225):689–97.

8. Volz E, Baguelin M, Bhatia S, Boonyasiri A, Cori A, Cucunubá Z, et al. Report 5: Phylogenetic analysis of SARS-CoV-2. 2020 [cited Apr 15, 2020]; Available from: https://www.imperial.ac.uk/media/imperial-college/medicine/sph/ide/gida-fellowships/Imperial-College-COVID19-phylogenetics-15-02-2020.pdf

9. Fang G, Li S, Liu Y, Xin N, Ma K. People beyond the statistics: Died of “normal pneumonia”? [统计数字之外的人:他们死于 “普通肺炎”?]. Caijing Zachi [财经杂志] (Finance and Ecomomics Magazine) 2020 [cited Feb 13, 2020]; Available from: https://web.archive.org/web/20200213190623/ http://www.sohu.com/a/370032279_120094087

## References

1. Anderson RM, May RM. Infectious diseases of humans. Oxford: Oxford University Press; 1991.

2. Drake JM, Bakach I, Just MR, O’Regan SM, Gambhir M, Fung IC-H. Transmission Models of Historical Ebola Outbreaks. Emerging Infectious Disease journal. 2015 Aug;21(8):1447–50.

3. National Health Commission of the People’s Republic of China. 2020 [cited Feb 2, 2020]; Available from: http://www.nhc.gov.cn/

4. Centre for Health Protection, Department of Health, The Government for the Hong Kong Special Administrative Region. 2020 [cited Feb 2, 2020]; Available from: https://www.chp.gov.hk/en/index.html

5. John Hopkins University Center for Systems Science and Engineering. 2019 Novel Coronavirus COVID-19 (2019-nCoV) Data Repository by Johns Hopkins CSSE. 2020 [cited Feb 13, 2020]; Available from: https://github.com/CSSEGISandData/COVID-19

6. Banks HT, Hu S, Thompson WC. Modeling and inverse problems in the presence of uncertainty: CRC Press; 2014.

7. Chowell G, Ammon CE, Hengartner NW, Hyman JM. Transmission dynamics of the great influenza pandemic of 1918 in Geneva, Switzerland: Assessing the effects of hypothetical interventions. J Theor Biol. 2006 Jul 21;241(2):193–204.

8. Chowell G, Shim E, Brauer F, Diaz-Duenas P, Hyman JM, Castillo-Chavez C. Modelling the transmission dynamics of acute haemorrhagic conjunctivitis: application to the 2003 outbreak in Mexico. Stat Med. 2006 Jun 15;25(11):1840–57.

9. Vynnycky E, White RG. An Introduction to Infectious Disease Modelling. Oxford: Oxford University Press; 2010.

